# Distinct cellular dynamics associated with response to CAR-T therapy for refractory B-cell lymphoma

**DOI:** 10.1101/2022.04.04.22273422

**Authors:** Nicholas J. Haradhvala, Mark B. Leick, Katie Maurer, Satyen H. Gohil, Rebecca C. Larson, Ning Yao, Kathleen M. E. Gallagher, Katelin Katsis, Matthew J. Frigault, Jackson Southard, Shuqiang Li, Michael C. Kann, Harrison Silva, Max Jan, Kahn Rhrissorrakrai, Filippo Utro, Chaya Levovitz, Raquel A. Jacobs, Kara Slowik, Brian P. Danysh, Kenneth J. Livak, Laxmi Parida, Judith Ferry, Caron Jacobson, Catherine J. Wu, Gad Getz, Marcela V. Maus

**Author notes:** These first authors contributed equally (Nicholas J. Haradhvala, Mark B. Leick, Katie Maurer, Satyen H. Gohil). These senior authors contributed equally (Catherine J. Wu, Gad Getz, Marcela V. Maus).

## Abstract

Chimeric Antigen Receptor (CAR)-T cell therapy has revolutionized the treatment of hematologic malignancies. Approximately half of patients with refractory large B-cell lymphomas achieve durable responses from CD19-targeting CAR-T treatment; however, failure mechanisms are identified in only a fraction of cases. To gain novel insights into the basis of clinical response, we performed single-cell transcriptome sequencing of 105 pre- and post-treatment peripheral blood mononuclear cell samples, and infusion products collected from 32 individuals with high-grade B cell lymphoma treated with either of two CD19 CAR-T products: axicabtagene ciloleucel (axi-cel) or tisagenlecleucel (tisa-cel). Expansion of proliferative memory-like CD8 clones was a hallmark of tisa-cel response, whereas axi-cel responders displayed more heterogeneous populations. Elevations in CAR-T regulatory cells (CAR-Tregs) among non-responders to axi-cel were detected, and these populations were capable of suppressing conventional CAR-T cell expansion and driving late relapses in an *in vivo* model. Our analyses reveal the temporal dynamics of effective responses to CAR-T therapy, the distinct molecular phenotypes of CAR-T cells with differing designs, and the capacity for even small increases in CAR-Tregs to drive relapse.

## Main text

CAR-T cell therapy has revolutionized the treatment of hematologic malignancies, resulting in multiple Food and Drug Administration (FDA) approvals since 2017. However, long-term responses only persist in roughly half of patients^1,2^. Investigations across hematologic malignancies have identified putative mechanisms for relapse including tumor resistance to apoptosis^3,4^, target antigen loss^5^, upregulation of inhibitory receptors, and intrinsic T-cell deficiencies^6,7^. For patients with chronic lymphocytic leukemia, a dearth of specific apheresis and CAR-T subpopulations appeared to be important for response^7^. However, thus far, investigation of mechanisms of resistance to CAR-T therapy in aggressive large-B cell lymphomas has been limited.

The first two FDA approved CAR-T cell products for large cell lymphoma, axicabtagene ciloleucel (axi-cel) and tisagenlecleucel (tisa-cel), utilize similar CAR designs with identical extracellular binding domains. They have, however, differing manufacturing processes and costimulatory domains (CD28 and 4-1BB respectively), endowing them with different expansion and phenotypic characteristics while maintaining comparable clinical efficacy^8^. A recent study of single-cell RNA sequencing (scRNA-seq) data on cellular infusion products from large cell lymphoma patients treated with axi-cel identified an association between complete clinical response and higher frequencies of CD8 T cells expressing memory signatures, while poor clinical response was associated with CD8 T cell exhaustion^9^. Much, however, remains to be understood with respect to how transcriptional mechanisms of response vary by product, evolve after infusion into the patient, or fit into the broader context of the host immune system.

We hypothesized that studying the evolutionary trajectories of patient-derived T cells and CAR-T cells from two different CAR products would yield insights into their respective modes of failure. Here, we performed scRNA-seq and single-cell TCR-sequencing of 105 samples from 32 patients with aggressive large B-cell lymphoma treated with axi-cel or tisa-cel, collected over the course of treatment. We found substantial differences in the cellular dynamics of response between the two products: tisa-cel responses were associated with striking expansion of rare CD8+ central-memory-like populations from the infusion products (IPs), while axi-cel treatment revealed less shifting of T-cell lineages between IP and post-treatment, and post-treatment cells in responders often originated from more-differentiated IP populations. In axi-cel IPs, we identified increased frequencies of CAR-Treg populations in patients lacking clinical responses. We then showed, through *in vitro* and *in vivo* modeling, that these CAR-Treg cells, expressing either product, can suppress conventional CAR-T activity and thus facilitate relapse.

## Results

To capture the evolution of CAR-T cells and other interacting immune cells, we performed scRNA-seq on peripheral blood mononuclear cells (PBMC) collected from multiple timepoints from 32 individuals with high-grade B-cell lymphoma treated with axi-cel (n=19) and tisa-cel (n=13), along with their matching infusion products (IP) (**Figs. 1a,b; Supplementary Table 1, 2**). Patient outcomes were tracked for up to six months, with fluorodeoxyglucose positron emission tomography (FDG-PET) scans taken for clinical response approximately 1, 3, and 6 months post-infusion for most patients. We observed 15 of 32 (47%) patients progress or relapse within the six-month window of observation, consistent with the known clinical activity of these agents (**Fig. 1b**). We analyzed PBMCs collected within 30 days prior to treatment (n=20) and, to capture the peak CAR-T expansion, also at 7 days after treatment (n=29). When possible, day 7 cells were sorted by flow cytometry based on expression of the CAR to generate an enriched fraction of CAR-T cells (n=22) (**Supplementary Fig. 1, Methods**). Altogether, our cohort yielded a total of 602,577 high-quality cells.

**Figure 1.**
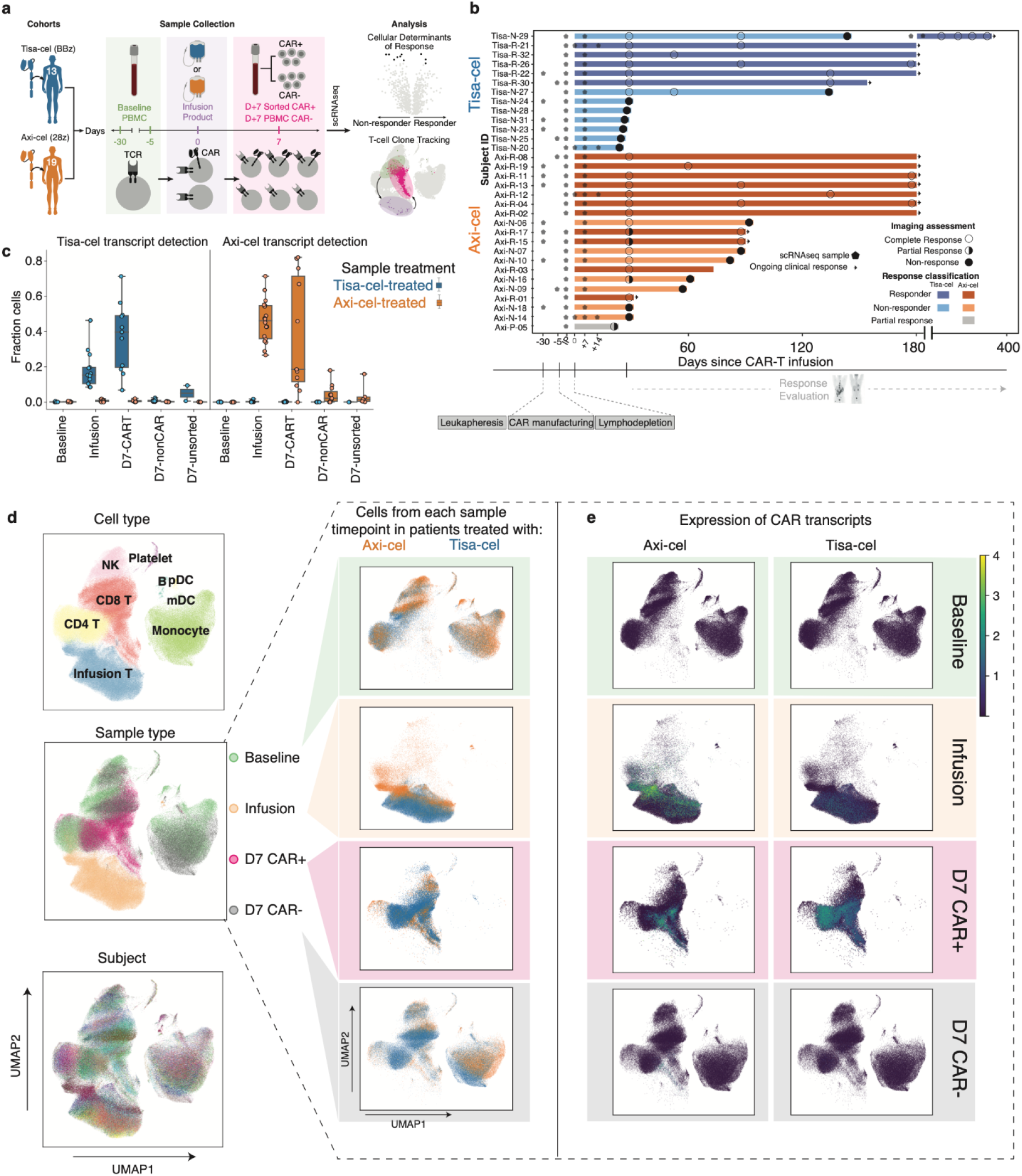
The landscape of CAR-T and host immune populations in CAR-T treated patients. **a**, Illustration of study design. **b**, Swimmer plot of patient outcomes. Bars represent follow-up window (either by PET scan or physician report), circles represent outcome of PET scans, and arrows represent ongoing response. Stars designate timepoints at which samples were collected for scRNA-sequencing. **c**, Fraction of cells per sample with at least one detected tisa-cel or axi-cel transcript. Each circle represents a sample, and samples from patients with tisa-cel or axi-cel are stratified. The timepoint and sorting strategy is denoted on the x-axis. **d**, UMAP representation of full dataset. On the left, colored by cell type, timepoint, and subject. On the right, cells from the same UMAP subset for each timepoint are colored by the CAR treatment (axi-cel orange, tisa-cel blue), and **e**, colored by the expression of each CAR construct detected in each cell.

To identify the CAR-transduced T cells based on transcript-level data, we leveraged *in-silico* detection of the respective CAR transcript (**Methods**). Cells with a detected CAR transcript were present at high numbers in the infusion products and the sorted CAR+ day 7 samples, while they were only minimally detected in the sorted CAR-negative fractions and unsorted samples from day 7 post-treatment samples (**Fig. 1c**). To assess the level of false CAR detection, we analyzed pre-treatment samples and artifactually detected the CAR transcript in only 73 of 131,000 cells (0.0006%).

Clustering of all cells in our dataset enabled the identification of diverse immune cell types present based on canonical markers **(Fig. 1d, Supplementary Fig. 2a**). Such analysis revealed day 7 non-CAR-T cells to intersperse with those from baseline, whereas day 7 CAR+ cells tended to cluster separately from the baseline and the day 7 CAR-negative cells, and infusion products clustered independently (**Fig. 1d, left, center plot**). Most clusters, and in particular the non-T cell clusters, spanned the patients, indicating that there was less variability among patients than among types of cells (**Fig. 1d left, bottom plot**). By segregating the data based on sample type (baseline, IP, day 7 flow-sorted CAR+ and CAR–), we further observed the absence of detectable CAR transcripts at baseline, rare CAR transcript expression in the day 7 CAR-negative sorted samples, and clustering of the CAR+ cells originating from the IP and from the CAR+ sorted samples (**Fig. 1e**). As expected, the IP samples and day 7 CAR+ samples primarily consisted of T cells, while baseline and day 7 CAR– samples contained other populations such as macrophages, myeloid, dendritic cells, and B cells (**Fig. 1d, right**). We did not detect major transcriptional differences among cells from patients treated with tisa-cel vs axi-cel **(Fig. 1d)**. However, cells from the IPs and day 7 samples appeared to cluster by CAR construct and expression, suggesting potential CAR-driven differences in their phenotype **(Fig. 1e)**.

### Transcriptomic evolution of axi-cel and tisa-cel from infusion to day 7

To molecularly identify the basis for the observed separation between transduced CAR-T cells and untransduced T cells, we devised a pseudobulk differential expression approach comparing cells of each category (**Fig. 2a, Supplementary Fig. 3, Methods**). At day 7, we found 463 genes to be elevated (FDR<10%, >2-fold increase in CAR+ cells relative to CAR-) in both types of CAR-transduced T cells (**Supplementary Table 3**). Many of the common top genes were related to cellular proliferation (*DIAPH3, BUB1B, KIF23*, q<10^−5^ for both products) and activation (e.g., *CD109* with q=2.1×10^−7^ and 1.0×10^−7^ for tisa- and axi-cel, respectively). Additional genes were found in product-specific analyses of both treatments. In the infusion products, axi-cel CAR+ cells had increased expression of *CTLA4* (q=5.0×10^−5^) and *STING1* (q=3.1×10^−4^) (**Fig. 2b**).

**Figure 2.**
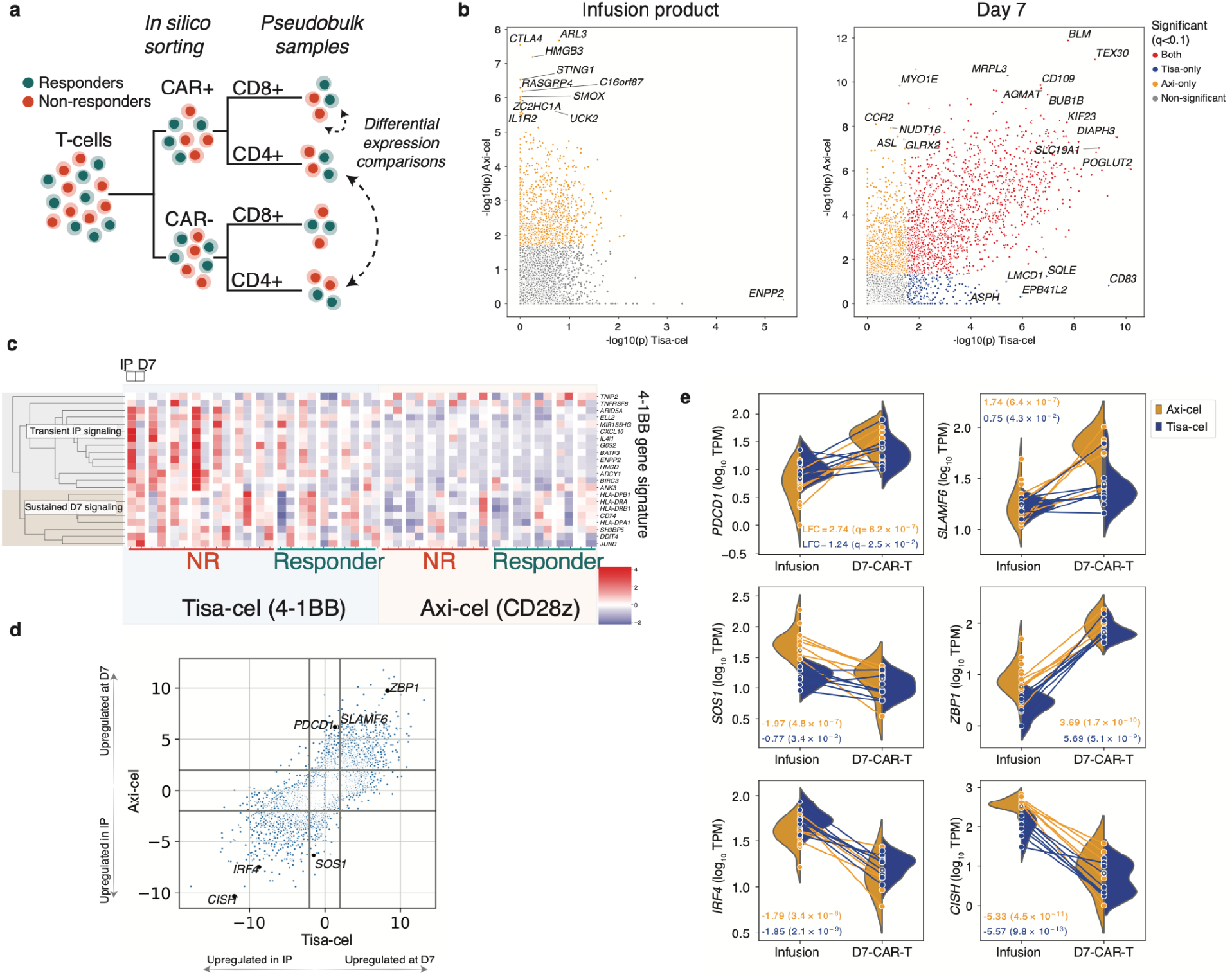
Pseudobulk analysis of genes related to treatment, timepoint, and response. **a**, Schematic of pseudobulk approach. T-cells from IP and day 7 samples are *in-silico* sorted for CAR+ and CAR-cells, and when stated CD4 and CD8. Transcript counts are then combined across these cells into one pseudobulk observation. Different conditions are then compared using limma^49^. **b**, Scatterplot of -log_10_ p-values from limma differential expression test between CAR+ and CAR- (not separating CD4 and CD8) pseudobulk samples from the IP (left, n=13 and n=18 for tisa-cel and axi-cel, respectively) and day 7 PBMCs (right, n=13 and n=15) for each product. **c**, Expression of previously identified signature of 4-1BB CAR activation across timepoints and patients. **d**, Comparison of genes differentially expressed between CD8+ cells in the infusion products (n=7 and n=15 for tisa-cel and axi-cel, respectively) and at day 7 (n=10 and n=7 for tisa-cel and axi-cel). The scatter plot shows the signed log_10_ p-values obtained from limma testing differences between IP and day 7 samples for tisa-cel (x-axis) and axi-cel (y-axis). Positive values indicate higher expression at day 7, and negative values higher in the IP. **e**, Illustration of several highlighted genes in panel d. Lines represent the changes in expression of the gene between infusion product and day 7 in CD8+ CAR-T cells of a single sample. Samples are colored by treatment, and split violins represent the distribution of expression across patients of each treatment at a given timepoint.

Our analysis of tisa-cel IPs yielded just one significant hit (q< 0.1), likely due to power limitations from the far lower CAR transcript detection rate compared to axi-cel **(Fig. 1c, Fig. 2b)**. This top differentially expressed gene, *ENPP2* (q=0.02), is associated with a 4-1BB signature^10^. Indeed we found genes of this signature to be recurrently upregulated in tisa-cel IPs over axi-cel (**Fig. 2c**). However we observed divergent temporal dynamics of the genes in this set, with a subset maintaining expression at day 7 (genes in the MHC II pathway), while others (such as the transcription factor *BATF3*) were transiently expressed in the IP but downregulated post-transfusion. To further map out these temporal transcriptional changes, we calculated differentially expressed genes between IP and day 7 for each product separately, and examined genes which were either commonly altered between the two products, or specific to a single product (**Fig. 2d, Supplementary Table 4**).

Some of the product-specific upregulated genes were expected, based on the known differences in CAR-T biology between CD28 and 4-1BB signaling in tisa-cel and axi-cel. *PDCD1* (PD-1), a CAR-T activation marker at early time points, was one of the most differentially upregulated genes at day 7 in axi-cel (log_2_ fold-change 2.74 day 7 relative to IP, q=6.2×10^−7^), a much stronger effect than seen in tisa-cel (log_2_ fold-change 1.24, q=2.5×10^−2^), aligning with the more rapid activation seen with CD28 signaling (**Fig. 2e**)^11–13^. Additionally, the immune checkpoint regulator *SLAMF6*, also associated with PD-1 elevation, was elevated in axi-cel at day 7 (log_2_ fold-change 1.74, q=6.4×10^−7^) and is known to play a role in T-cell exhaustion and impaired effector function and may be suggestive of an early exhaustion signature in this rapidly expanding product^14^. Further supporting the rapid kinetics of CD28 signaling was upregulation of *SOS1* in the IP of axi-cel, which is associated with CAR proliferation^15^ (log_2_ fold-change −1.97, q=4.8×10^−7^).

We also identified 1442 differentially expressed genes shared between the products (708 upregulated and 734 downregulated by at least 2-fold in day 7 samples compared to IP with q<0.1). One of the genes most significantly upregulated in both products was *ZBP1* (tisa-cel q=5.1×10^−9^ and axi-cel q=1.7×10^−10^) which plays an important role in mediating necroptosis in response to interferon-exposed cells^16^, suggestive of potential upregulation of activation-induced cell death pathways in both products at this timepoint. *ZBP1*-deficient mice resist systemic inflammation driven by elevations in IFNg signaling, suggesting a potential pathway for regulation of CAR toxicity^16,17^. Two genes were notable when examining the top 10 genes with the most significantly lowered expression at day 7 compared to the infusion product in both tisa-cel and axi-cel: (i) *IRF4* (tisa-cel q=2.0×10^−9^ and axi-cel 1=3.4×10^−8^) has been implicated in prevention of exhaustion in the CAR-T cells of *in vivo* cancer models^18^; and (ii) *CISH* (tisa-cel q=9.8×10^−13^ and 1=4.5×10^−11^) which has been identified as a critical T-cell immune checkpoint, the ablation of which results in enhanced T-cell function mediated through the TCR^19^.

### Associations of PBMC and CAR-T cell subsets with clinical response

We next queried our dataset for cell types with frequencies associated with clinical outcomes. Patients were binarily categorized into responders and non-responders based on whether progressive disease was observed by PET within six months of follow-up (**Methods**). Among the CAR-negative PBMCs, we used the method scCODA^20^ to test whether any non-T-cell populations changed in frequency relative to the number of T-cells. We identified monocytes as more abundant in non-responders at baseline for both products (FDR=0.09, estimated log_2_ fold-change of 0.76±0.38 between non-responders and responders, **Supplementary Fig. 2b**), in agreement with previously described results^21^. This effect was not observed at day 7, nor were any other non-T cell populations found to be significantly altered.

Next, we focused on differences in the T cell compartments. We first assessed whether the relative fractions of CD4+ and CD8+ subsets were associated with response (**Supplementary Fig. 2c**). For patients treated with tisa-cel, but not axi-cel, response was associated with the extent to which day 7 CAR+ cells were predominantly (>50%) of CD8+ subtype (**Fig. 3a;** likelihood ratio test p=0.02 and p=0.51 for tisa-cel and axi-cel, respectively). This trend was captured by the categorization of response based on a 6-month window of observation (**Supplementary Fig. 2c,d**; BH-corrected two-tailed t-test q=0.054), and not a shorter 1-month window (q=0.78). Five of the seven tisa-cel treated patients with lower proportion (<50%) of CD8+ CAR+ T cells at day 7 exhibited progressive disease by day 30, and the remaining two relapsed six months following treatment. In contrast, only one patient (Tisa-N-31) of the six patients with high proportion of CD8+ CAR-T cells at day 7 exhibited progression of disease. In addition, the percentage of CD8+ cells in tisa-cel infusion products was low in both responders and non-responders, but had a modest association with 6-month response (**Supplementary Fig. 2c;** q=0.11).

**Figure 3.**
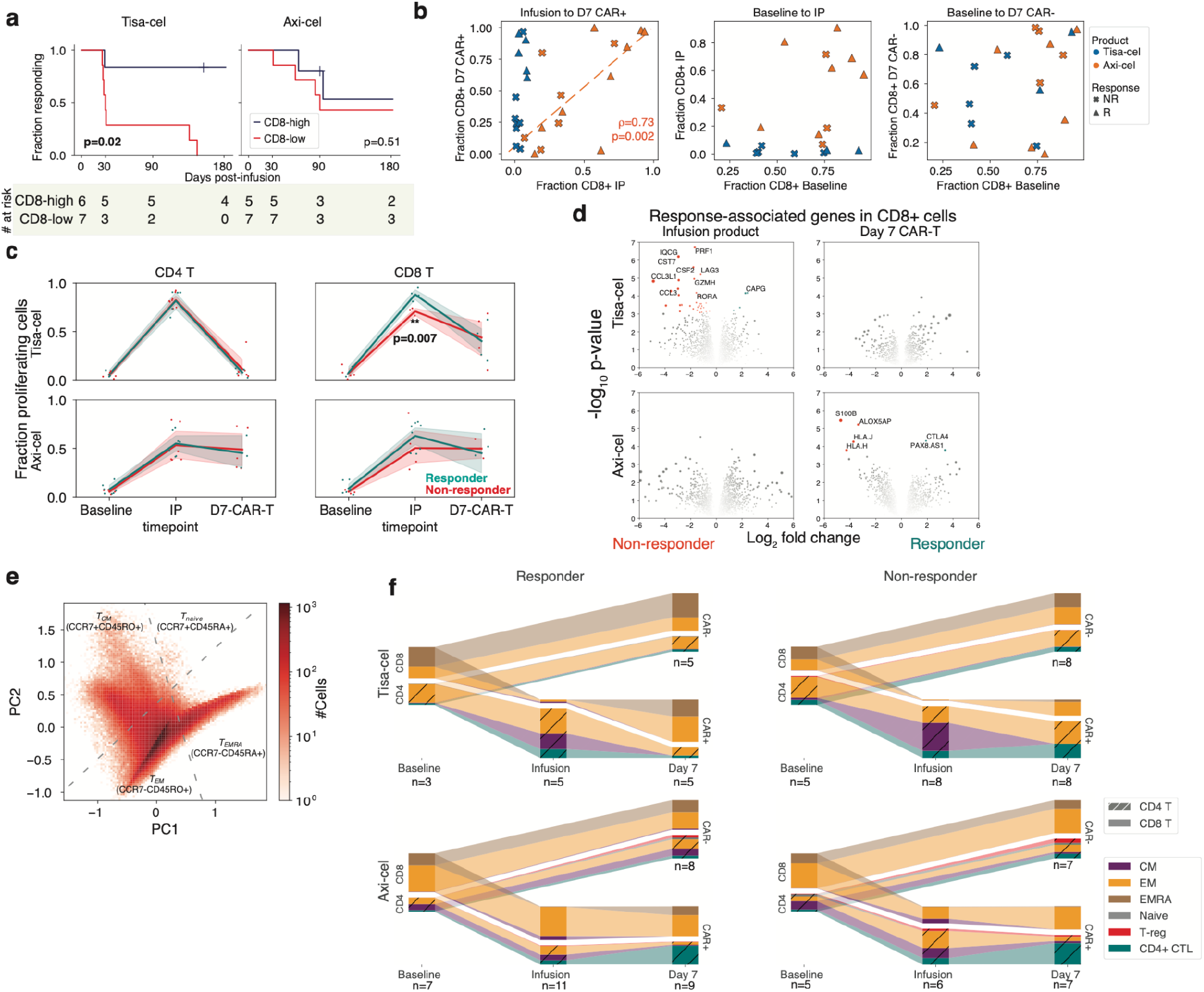
Temporal evolution of T cells in responders and non-responders of different products. **a**, Kaplan-meier response curves stratified by whether patients had >50% CD8 CAR-T cells (out of all CAR-T cells) at day 7. P-values testing the significance of this stratification using a cox proportional-hazards model are shown. **b**, Changes in CD8 frequency between different timepoints for CAR- and CAR+ cells. **c**, Fraction of cycling cells at different timepoints for different CD4/8 designations, treatments, and response outcomes. A two-tailed t-test is shown for tisa-cel CD8+ IPs comparing responders and non-responders (p=0.007, t-statistic=-4.3, n=4 and n=3 for R and NR). Individual samples are shown as dots, and the band represents the standard deviation. **d**, Volcano plots of differentially expressed genes in CD8+ CAR-T cells between responders and non-responders for each product and timepoint. All genes with Benjamini-Hochberg FDR-corrected p-values (as determined by limma) are colored and labeled. The area of each dot is the absolute value of the log_2_ fold change times the negative log_10_ p-value. **e**, PCA dimensionality reduction using knn-smoothed expression of T differentiation markers (*CD45RO, CD45RA*, and *CCR7*) visualizing the classification of T cell subtypes. Dotted lines represent where the cutoffs used to define *CCR7+* and *CD45RA+* (smoothed expression values >0.5) fall in the projection. **f**, Depiction of T subset frequencies at each timepoint for each product and response. Bar widths at each timepoint are proportional to the fraction of cells (out of all calls) being classified as a particular subset. CD4 and CD8 cells are stratified, and distinguished with cross-hatching for CD4 subsets.

The temporal dynamics of these CD8 fractions from IP to day 7 were strikingly different between the two treatments. In axi-cel, the fraction of day 7 CAR+ T cells that were CD8+ was highly variable, and this value was highly correlated with the percentage of CD8+ cells in the infusion product (**Fig. 3b, left** spearman ***ϱ***=0.73, p=0.002). Tisa-cel IPs, however, were universally composed of <10% CD8+ cells, but these CD8+ cells dramatically expanded at day 7, particularly in the patients exhibiting long-term response (**Fig. 3b, left**). Baseline fractions of CD8+ cells were found to be predictive of neither those observed in the circulating CAR-negative T cell populations nor in the IPs (**Fig. 3b, center and right, respectively**).

### Distinct transcriptional features underlie CD8+ T cell expansion and response between the two products

By scoring our cells for cell cycle gene sets^22^, and calculating the fraction of cycling CAR-T cells at each timepoint, we found that the different dynamics of CD4+ and CD8+ T cells could be explained by product-specific proliferation behaviors (**Fig. 3c)**. Tisa-cel CD4+ CAR-T cells exhibited cycling rates comparable with CD8+ cells in the IP but essentially ceased all proliferation at day 7, explaining the outgrowth of CD8+ cells. Responders had higher CD8+ proliferation rates in the IPs (two-tailed t-test p=0.007), likely underlying the greater expansion seen at day 7. Meanwhile, axi-cel CD4+ CAR-T cells exhibited similar proliferation rates to CD8+ cells both in the IP and at day 7, consistent with them maintaining similar ratios across timepoints.

By testing for genes differentially expressed between responders and non-responders in our pseudobulk framework, at each timepoint and for CD4+ and CD8+ cells separately, we found that the majority of differentially expressed genes (14/17 genes with FDR<10%) were in the CD8+ cells of tisa-cel infusion products (**Fig. 3d, Supplementary Table 5**). Non-responders showed a clear activated cytotoxic phenotype, with upregulation of genes such as perforin (*PRF1*), *GZMH*, and *LAG3*. Thus, further differentiation of these cells provides a possible explanation for the lower proliferation and failure of expansion in non-responders. We did not observe differential expression when studying CD8+ axi-cel infusion products. Prior work^9^ studying axi-cel infusion products had proposed dysfunctional and memory genes characteristic of non-responders and responders respectively, but examining these genes in our data did not yield a significant separation of patients by response (**Supplementary Fig. 4**).

To better understand our signature of non-response in tisa-cel infusion products, we mapped the cells onto canonical T cell subsets on the basis of conventional flow cytometry based definitions. We developed an Expectation-Maximization-based method to estimate *CD45RA* and *CD45RO* isoform expression from the single cell transcriptome data (**Supplementary Fig. 5a,b, Methods**). We then used smoothed expression of common T-cell subset markers (**Methods, Supplementary Table 6)** to classify the cells into naive (*CCR7+CD45RA+*), central memory (CM, *CCR7+CD45RO+*), effector memory (EM, *CCR7-CD45RO+*), terminally differentiated effector memory re-expressing *CD45RA* (EMRA, *CCR7*-*CD45RA*+), and regulatory T-cells (*FOXP3*+) (**Fig. 3e, Supplementary Fig. 5c-f**). We further distinguished CD4+ cytotoxic cells (CD4+ CTLs, *NKG7*+), prevalent in our dataset, from other EMs with a more typical helper phenotype (**Supplementary Fig. 5d,f**).

Tisa-cel IPs were composed primarily of CD4+ cells with a mix of CM and EM phenotypes, with a modest elevation of CMs in non-responders compared to responders (**Fig. 3f**; median of 24% and 56% cells for responders and non-responders, respectively; two-sided t-test p=0.084). At day 7 the CM phenotypes were absent, having been replaced by EM populations, including CD4+ CTLs in 3 of 8 of the non-responding patients (and seen in 0 of 5 of responders). The previously noted post-treatment expansion of CD8+ cells in tisa-cel took on a primarily *CCR7-CD45RO+* EM phenotype regardless of response, with cells beginning to re-express *CD45RA* (EMRA) in several patients (**Supplementary Fig. 5g**). In axi-cel-treated patients, an expansion of CD4+ CTLs was also seen following treatment (median of 9.2% and 30% of cells in IP and day 7 CAR-T populations respectively, t-test p=0.0032) in responders and non-responders alike. CAR-negative cells in patients treated with either treatment were a mix of EM and EMRA phenotypes which remained unchanged in frequency from baseline to post-treatment (**Fig. 3f**).

To further dissect these populations in an unsupervised manner, we performed clustering of IP and day 7 CAR-T cells for each treatment and clinical outcome (**Fig. 4a, Supplementary Fig. 6a**). CD8+ tisa-cel IP cells demonstrated a considerable shift in their distribution across the identified clusters when comparing responders and non-responders (**Fig. 4b, Methods**, log likelihood ratio 20.3, BH-corrected q=0.060). Tisa-cel responders had more cells from the cluster expressing CM markers *CCR7* and *LEF1* (two-tailed t-test p=0.0057, **Fig. 4c,d**). They also expressed the transcription factor *STAT1*. Non-responders had higher frequencies of cells falling into cluster EM 1 (two-tailed t-test p=0.0062) with markers similar to those observed in our pseudobulk analysis: interferon gamma (*IFNG*), cytotoxicity (*PRF1*), and markers of activation/exhaustion *LAG3* and *HAVCR2* (TIM-3) (**Fig. 4c,d**). While overall a shift in the subtypes of CD8+ tisa-cel CAR-Ts at day 7 was not observed (log likelihood ratio 0.34, q=0.98), we found that the only non-responder that had >50% CD8+ cells out of their CAR-T cells post-treatment (Tisa-N-31) had a distinct phenotype from the responders in our dataset (**Fig. 4e,f**). Their cells were distinguished by high expression of *KLRG1* and lack of L-selectin (*SELL)*, a phenotype attributed in the literature to short lived effector cells (SLECs)^23^. Thus despite the CD8+ expansion in this patient, these cells may not have had the correct phenotype to induce an effective response. The cells of most responders fell into an *IL7R*+ EM 1 cluster, with the exception of one patient (Tisa-R-22) with a unique *CX3CR1*+*CD27*+ phenotype (**Fig. 4e, Supplementary Fig. 6b**, cluster EM 2).

**Figure 4.**
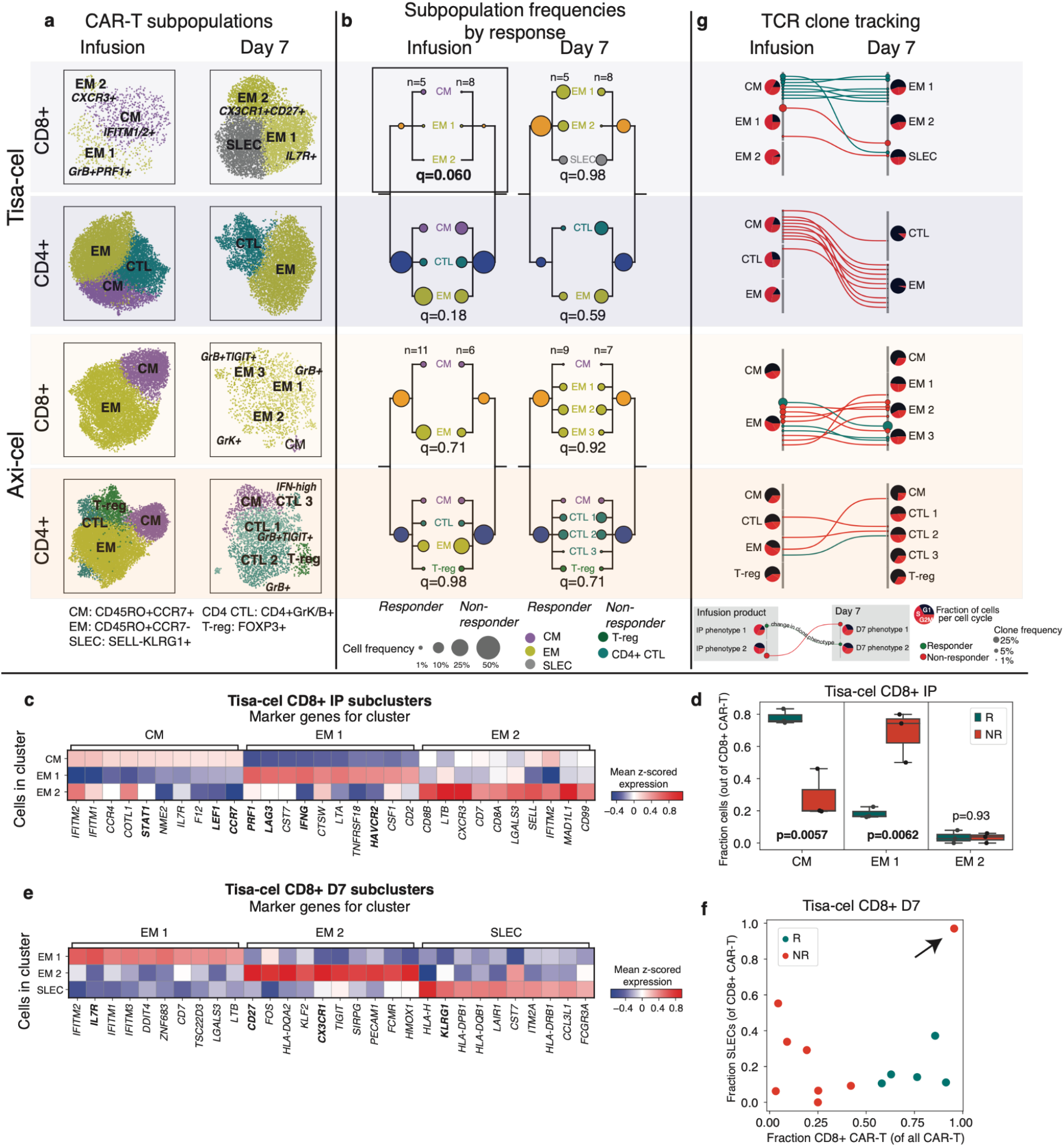
Tracking temporal evolution of CAR-T clones by TCR sequencing. **a**, u-map plots of CAR-T cells of different treatments and at different timepoints colored by subcluster. **b**, Frequencies of CAR-T subclusters. Circle area represents the estimated median percentage of cells belonging to the cluster (out of all CAR-T cells) in patients treated with each product and with each response outcome. **c**, Top 10 differentially expressed genes in each tisa-cel CD8+ IP subcluster, as identified by a t-test. **d**, Relative frequencies of each tisa-cel CD8+ IP cluster stratified by response. Only samples with at least 25 CD8+ CAR-T cells are shown. **e**, Top differentially expressed genes in each tisa-cel CD8+ day 7 subcluster, as identified by a t-test. **f**, Scatter plot of the fraction of CD8+ tisa-cel day 7 cells in each patient that fell into the SLEC cluster (y-axis) vs the overall fraction of CAR-T cells that were CD8+ (x-axis). Arrow highlights the sole non-responder to have predominantly CD8+ cells are day 7. **g**, Up to 15 of the most prevalent TCR clones identified at both time points are shown for each CAR-T subset. For each, circles show the cluster in Figure 3D to which the clone belongs at each timepoint, with sizes corresponding to the clone frequency in its sample. Pie charts show the distribution of cells in each phase of the cell cycle.

We next used our TCR enrichment libraries to track the temporal evolution of the CAR-T phenotypes. In all tisa-cel responders (and several non-responders), the top CD8+ TCR clones in the IP were found to be increased in frequency at day 7 (**Supplementary Fig. 7**). In responders the cluster membership of these clones showed that they primarily originated from CMs in the IP and differentiated into *IL7R*+ EMs at day 7 **(Fig. 4g)**. Meanwhile clones with a SLEC phenotype at day 7 were inferred to have differentiated from the non-response-associated *GZMB*+*PRF1*+ EM population. Axi-cel again showed starkly different dynamics, where CM phenotype cells in the IPs exhibited lower rates of proliferation than other IP subsets, and were not observed to be the primary origin of the CAR-T cells present at day 7 **(Fig. 4g)**, suggesting a less predominant role, at least initially, of these cells compared to tisa-cel.

Finally, we identified a population of regulatory T cells with CAR transcripts present (CAR-Tregs) in the infusion products. Tregs represent a rare subset of CD4+ circulating lymphocytes that play a critical role in preventing autoimmunity^24^. Tregs are also implicated as tumor-cooperating partners in the hostile, immunosuppressive tumor microenvironment in a variety of tumor types, including aggressive lymphomas^25^. *In vivo* models of high proportions of CAR-Tregs have been shown to be potently suppressive in both cancer and organ transplant models ^26–29^, but CAR-Tregs have not yet been shown to play a role in clinical relapse after CAR T cell treatment.

While CAR-Tregs were identified initially in axi-cel products (**Fig. 4a**), by analyzing tisa-cel and axi-cel together with batch correction for inter-product differences, we also identified CAR+CD3+*FOXP3*-high cells in tisa-cel IPs, albeit at substantially lower frequencies than seen in axi-cel (**Fig. 5a**, median 0.7% of all CAR+ cells in tisa-cel vs. 3.4% in axi-cel, two-tailed t-test p=9.7×10^−5^). We found that in axi-cel IPs this population was increased in non-responders (two-tailed t-test p=0.011), with an increased but non-significant trend in tisa-cel non-responders as well (two-tailed t-test p=0.53). The frequency of CAR-Tregs (of the total CAR+ T cells) in the axi-cel infusion products in our cohort was only a median of 5.6% and 2.8% (with ranges of 3-8% and 0-4%) in non-responders and responders, respectively. In exploring an external single-cell data set^9^ of axi-cel infusion products, we also found a trend towards higher T-regulatory cells in non-responders (**Fig. 5b**, two-tailed t-test p=0.17). Most striking were several nonresponders with very high CAR-Treg frequencies, up to 26.7% of all CAR+ cells (range 4.5-10.3% for responders, and 3.7-26.7% for non-responders).

**Figure 5.**
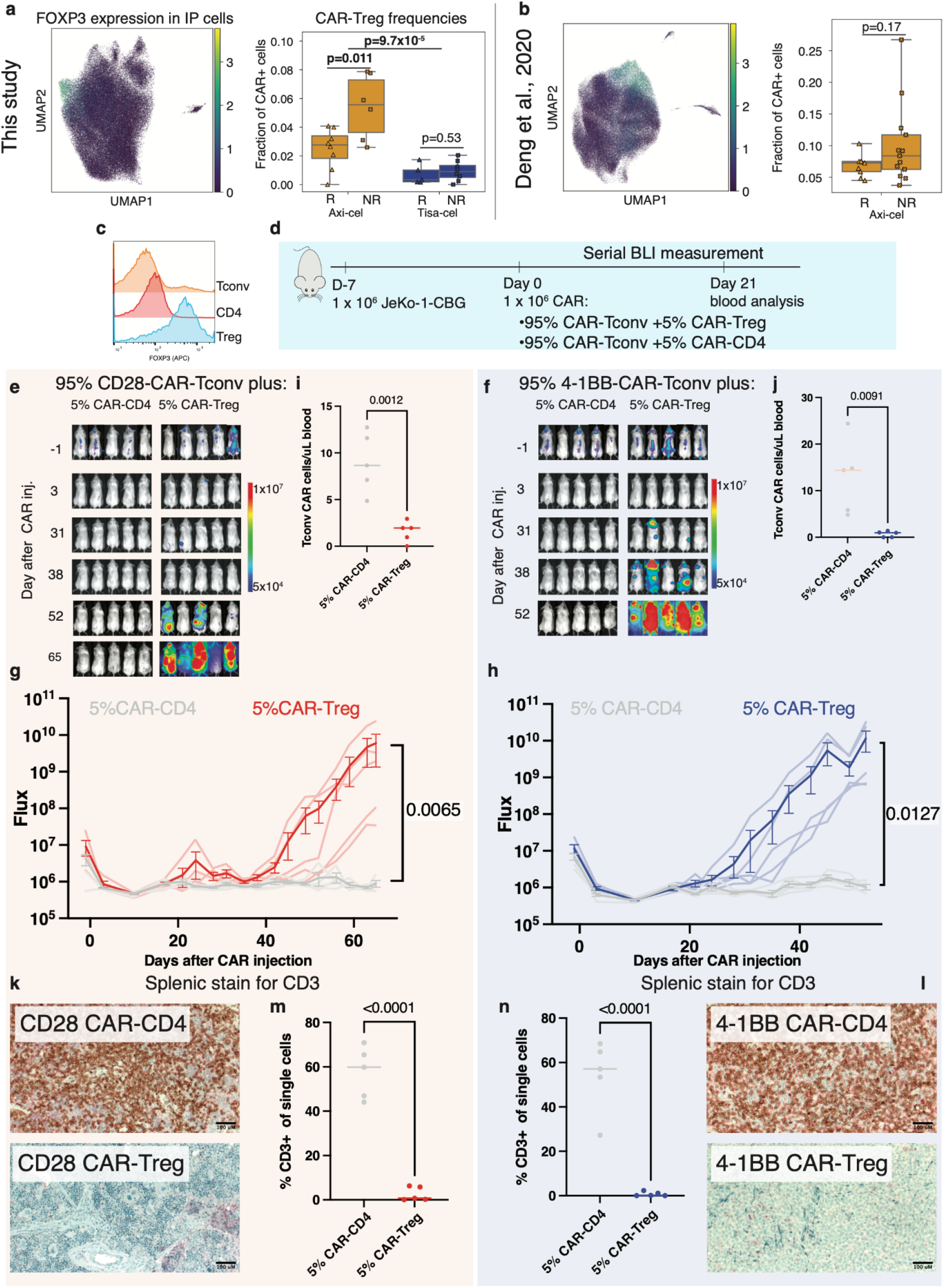
Mediation of relapse by CAR-Tregs in patients and *in vivo* validation experiments. **a**, Summary of CAR-Treg representation in this dataset as well as **b**, an independent set of axi-cel infusion products. Shown are a u-map representation of cells colored by FOXP3 expression (left) and the fraction of identified CAR-Tregs (out of all CAR-T cells) stratified by product and response (right). **c**, Intracellular flow cytometry staining of FOXP3 staining of donor T-regs, CD4 control T-cells, and Tconv T-cells. **d**, Schematic of *in vivo* CAR-Treg validation. NSG mice were injected with 1 × 10^6^ Jeko-CBG lymphoma cells on day −7. On day 0 mice were injected with 1 × 10^6^ CAR-T cells representing 95% CAR-Tconvs with either 5% CAR-Tregs or 5% CD4-CAR-T control cells. Experiment performed with CD19-CD28 (left) or CD19-4-1BB (right) constructs. **e,f**, Time course tumor radiance (photons/sec/cm2/sr) and **g,h**, flux (photons/s) for CD28 and 4-1BB experiments respectively. Mean ± SEM overlayed on individual subject curves. P-value represents the result of two-way ANOVA. **i,j**, Flow cytometric quantification of CAR-Tconv day 14 after CAR injection. P-value represents two-tailed unpaired t-test. **k,l**, Representative immunohistochemical staining for human CD3 in the spleen. **m,n**, Flow cytometric quantification of CD3 cells from the spleens of the indicated conditions. P-value represents two-tailed unpaired t-test.

### Nominal increases of infused CAR-Tregs are sufficient to drive lymphoma relapse *in vivo* in aCD19-28 and aCD19-BB treated mice

To assess whether CAR-Tregs could drive resistance in aggressive lymphomas at the proportions observed in the infusion products, we pursued *in vitro* and *in vivo* modeling. We isolated bulk CD3+ conventional T-cells (Tconv), T-regs, and CD4+ T-cells from healthy donor peripheral blood, and transduced these populations with either CD28 or 4-1BB costimulated second-generation CD19-targeted CAR constructs (**Fig. 5c, Supplementary Fig. 8a-b**). By CFSE staining of the bulk population, a population of 25% CAR-Tregs reduced CAR-Tconv expansion compared to CAR-CD4 controls for both constructs (**Supplementary Fig. 8c,d**). To assess the contribution of CAR-Tregs to relapse in an *in vivo* model of lymphoma, we subjected NSG mice engrafted with JeKo-1 lymphoma cells to injection of CAR-T-cells composed of 100% CAR-Tconv, or 75% CAR-Tconv with either 25% CAR-CD4 or 25% CAR-Tregs (**Methods, Supplementary Fig. 9a**). All mice appeared to rapidly clear tumor by day 3 (**Supplementary Fig. 9b,c**). However, for both constructs, the overall numbers of CAR-Tconv cells in the peripheral blood at day 14 were suppressed in the CAR-Treg treated mice (**Supplementary Fig. 9d,e**), and most of the mice treated with products containing CAR-Tregs relapsed, whereas none of the control mice had at the time of euthansia on day 35 (**Supplementary Fig. 9f,g**). Quantification of CAR-Tconv cells in the spleen at the time of euthenasia revealed high levels in the CAR-CD4 control mice and minimal levels of CAR-Tconv in the mice treated with injections containing CAR-Tregs (CD28 median 37.0 vs. 2.3, p=0.007; 4-1BB median 54.1vs 0.6, p=0.0003, **Supplementary Fig. 9h-k**).

After demonstrating that 25% CAR-Tregs were sufficient to drive relapse, we next wanted to ascertain if a lower, more physiologically relevant proportion of CAR-Tregs (as seen in our cohort) could also drive relapse. We engrafted NSG mice with JeKo-1 lymphoma cells followed by injection of CAR-T cells 7 days later, composed of 95% CAR-Tconv with either 5% CAR-CD4, or 5% CAR-Tregs (**Fig. 5d**). Again, after apparent early tumor clearance in all mice by day 3, all CAR-Treg mice developed frank relapse after extended follow up, while no relapses were seen in control mice (**Fig. 5e-h**). Assessment of peripheral blood CAR-Tconv expansion at day 21 revealed suppression in the CAR-Treg treated mice relative to control mice (CD28 median 8.7 vs 2.0, p=0.0012; 4-1BB 14.4 vs. 0.9 p=0.0091, **Fig. 5i,j**). At the time of euthansia, splenic CD3 staining was markedly reduced in all CAR-Treg mice by IHC and flow cytometry (CD28 median 59.8 vs. 0.68 p=<0.0001, 4-1BB median 57.1 vs. 0.12, p<0.0001, **Fig. 5k-n**). Thus, CAR-Tregs making up merely 5% of total infused CAR-T cells were sufficient to drive late tumor relapses and suppress CAR-Tconv expansion.

### Differential expansion of CAR-T populations between first and second treatments in association with clinical response

One of the study subjects (Tisa-N-29) that received tisa-cel had a CD19+ relapse 6 months after treatment. This patient was subsequently treated with a second infusion of the same infusion product and then achieved clinical and radiographic complete remission (last follow up 215 days after infusion; **Fig. 6a**).

**Figure 6.**
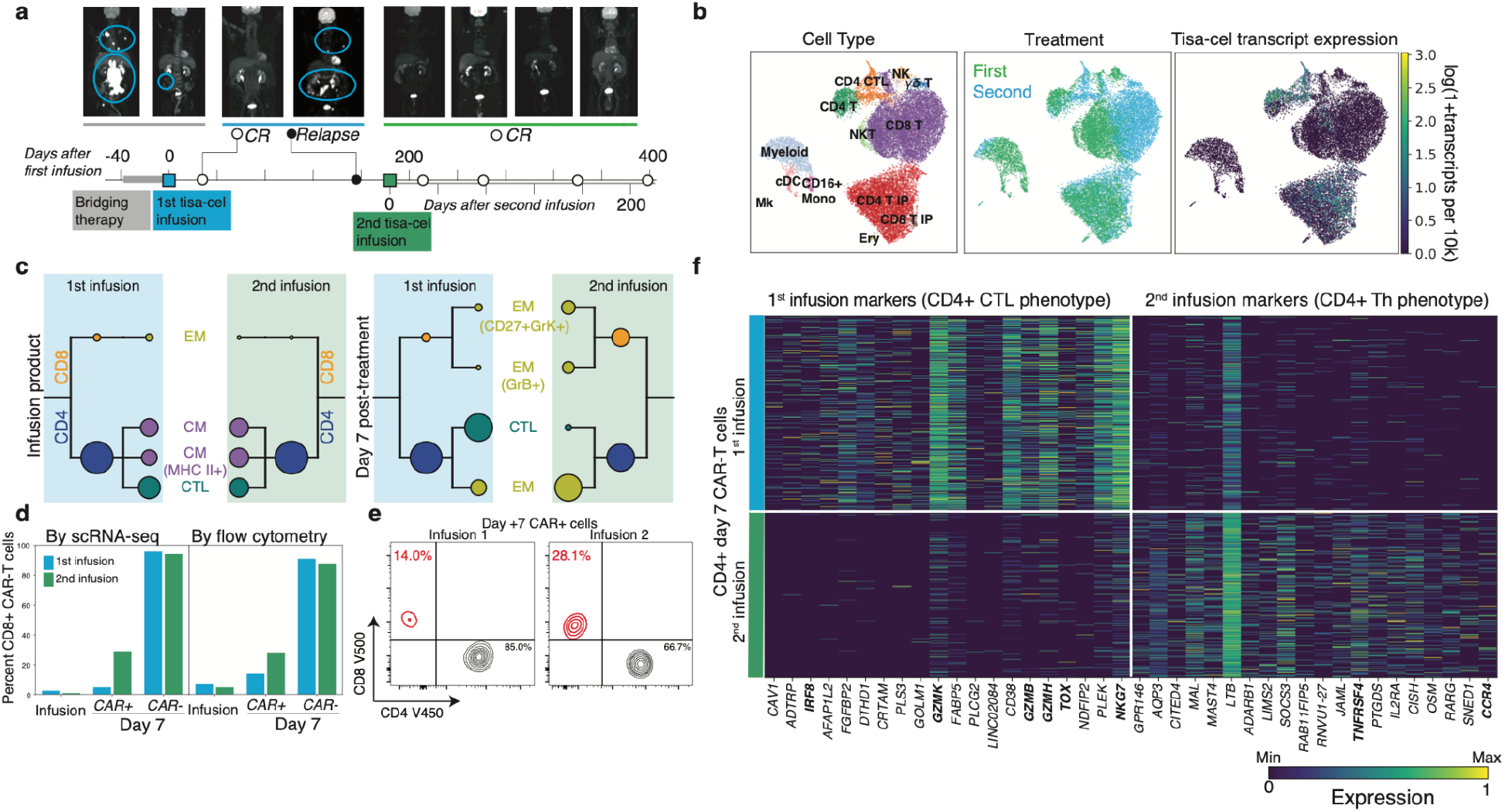
Differential characteristics of tisa-cel expansion in patient with relapse and subsequent retreatment. **a**, PET scans and illustration of treatment timeline for patient with relapse and subsequent re-treatment. **b**, U-map representation of cells from patient with second treatment colored by cell type, CAR (tisa-cel) expression, and first or second infusion. **c**, CAR-T subpopulation frequencies using the same visualization described in Fig. 4b. **d**, Fraction of CD8+ cells found by scRNA and flow-cytometry in different T cell subsets for each treatment. **e**, Flow cytometric measurements of CD8+ cell fractions in day 7 CAR-T cells. **f**, Top differentially expressed genes between the two treatments for CD4+ CAR-T cells, as determined by a t-test.

PBMCs collected seven days after re-treatment as well as the re-administered infusion product were analyzed by scRNA-seq as before (**Fig. 6b**). As expected, the two IP products lacked any discernible differences in cellular composition (**Fig. 6b,c**). In contrast, CAR-T cells at day 7 following each of the treatments exhibited marked differences. While few (5%) CD8+ CAR-T cells were detectable seven days after the initial infusion, ∼30% (a six fold increase) was observed after the second (**Fig. 6d**), a finding confirmed by parallel flow cytometry measurements (**Fig. 6d,e**). Between the two infusions, no differences in phenotype were found in the post-infusion circulating CD8+ CAR-T cells, although statistical power for this comparison was limited by the low frequency of CD8+ T cells after the first treatment. On the other hand, the phenotype of CD4+ CAR-T cells strikingly shifted from one treatment to the other (**Fig. 6c,f**). The CAR-T cells had a CTL phenotype after the first infusion, with top differentially expressed genes including markers of cytotoxicity (*GZMK, GZMB, GZMH, NGK7*), the transcription factor *IRF8*, and the gene *TOX*, implicated previously in T cell exhaustion^30^. In contrast, CD4+ cells after the second infusion instead expressed genes more consistent with a helper phenotype such as the co-stimulatory molecule OX40 (*TNFRSF4*) and Th2-marker *CCR4*.

Differences were also observed in the non-CAR compartment, with the first post-treatment sample being composed of almost exclusively CD8+ T cells, while the second additionally contained myeloid and NK cells (**Fig. 6b, Supplementary Fig. 10a, b**). The CD8+ non-CAR-T cells exhibited substantial differences in expression between the two timepoints. After the first treatment, these cells expressed *CD27* and *CXCR3*. After the second they expressed higher levels of transcription factors *GATA3* and *TBX21* (t-bet) and increased levels of *GNLY* (**Supplementary Fig. 10c**).

## Discussion

CD19 CAR-T therapy has provided a cure to roughly 40% of patients with previously incurable lymphomas^31,32^, but the failure modes in the remaining patients remain incompletely understood^33–35^. Collectively, our study represents the largest clinically-derived scRNAseq dataset of CAR-T cells and the circulating host immune cells assembled thus far and provides important observations of temporal changes in clonal and expression dynamics of the two major CD19 CAR-T designs in use for large-cell lymphoma (**Supplementary Fig. 11**). We find that tisa-cel responses are driven by expansion of CD8 populations with a central-memory phenotype, which are infrequent in the originating infusion product. In contrast, axi-cel products rely on pre-expanded populations with a further differentiated phenotype split more heterogeneously among CD4 and CD8 cells, and IP cells with a central memory phenotype that do not seem to expand at day 7 by TCR tracking.

A striking feature of our data is the divergent phenotypes and apparent response mechanisms of the two products. In addition to the differing co-stimulatory domains (4-1BB for tisa-cel, and CD28 for axi-cel), the products have other structural differences in the receptor and have important differences in the manufacturing process^36^, including how the initial apheresis product is prepared and shipped (fresh vs frozen) and how the activation is performed (antibody-coated beads vs soluble antibody and cytokines). Future work to pinpoint the mechanisms underlying the associations seen in this study will help illuminate alterations to the manufacturing process that may improve the efficacy of specific products. Another product, lisocabtagene maraleucel (liso-cel)^37^, gained FDA approval during the course of this study, and as it employs a 4-1BB costimulatory domain but is manufactured with a fixed CD4:CD8 ratio, future comparison of liso-cel and tisa-cel dynamics using the techniques presented here could be illuminating into the biological ramifications of a pre-defined ratio.

While tisa-cel IP phenotypes were found to be predictive of response, we also demonstrate a case where re-administration of a second infusion of the same IP to a patient resulted in expansion of different immune cell populations after each treatment. The fact that these two CAR-T infusions were drawn from the same manufacturing run highlights the importance of CAR-T-extrinsic phenotypes in response. How important is the host immunological milieu for CAR-T cell therapy and to what extent can we optimize it? Do the stochastics of infusing individual unique cells matter more than previously thought? Retrospective studies of patients who have received multiple infusions suggest that this successful re-induction of remission is rare^38^.

Finally, we found that increases in infusional CAR T-regulatory cells (CAR-Tregs) are associated with non-response to CD28-based (axi-cel) or 4-1BB-based (tisa-cel), and *in vivo* modeling suggests that this may be sufficient to drive late relapses and suppression of CAR-Tconv expansion in lymphoma. While the inhibitory effects of CAR-Tregs are well established, this is the first demonstration that small numbers of CAR-Tregs are capable of driving clinical relapse. This has important implications for the adoptive cellular therapy field. Depletion of T-regulatory cells from mobilized stem cell products using CD25 selection has been performed using GMP procedures and at clinical scale in the context of an autologous stem cell transplant for multiple myeloma^39^, suggesting that such a step could be added to traditional CAR-T manufacturing pipelines. Additionally, despite similar levels of CAR-Tregs in responding and non-responding patients who received tisa-cel, *in vivo* modeling demonstrated that CAR-Tregs incorporating either costimulatory domain could cause relapse. This seeming disparity could reflect the significantly lower number of CAR-Tregs in tisa-cel in our patient data, possibly a consequence of the different manufacturing procedures. Axi-cel manufacturing begins from fresh apheresis cells while tisa-cel is derived from frozen cells, and we speculate that cryopreservation may serve as a crude depletion of T-regs, which are notoriously intolerant of freezing^40–42^.

In summary, we have profiled the transcriptomic evolution of immune cells over the course of treatmenting refractory B-cell lymphomas with the first two CD19 CAR-T products. It is our hope that through illumination of the response-associated characteristics of each treatment we can optimize the design of CAR-T therapies, match patients with the treatments most likely to induce a clinical response, and understand strategies for combating relapse.

## Methods

### Patient samples and cell preparation

All patients were treated with commercial CAR-T cell products; clinical data and blood samples were obtained after written informed consent under an IRB-approved protocols at the Dana-Farber/Harvard Cancer Center (DFHCC #16-206 and 17-561). We complied with all relevant ethical regulations for human research participants. Responders were defined as those without radiographic relapse by 6 months of follow-up. Non-responders were defined as patients whose disease either did not have an initial response at day 28 or who progressed before the 6-month follow-up window. Infusion products were collected from the remnants of infusion bags and cryopreserved in a solution of 10% DMSO in FBS.

Cryopreserved PBMC from day 7 after CAR-T infusion were thawed and resuspended in FACS buffer (phosphate buffered saline (PBS) with 2% fetal bovine serum (FBS)). 5uL of Fc block (BD Bioscience) was added for 5 minutes followed by addition of the antibodies in BD brilliant stain buffer (BD Bioscience). The cells were stained for 20 minutes at 4°C using the following antibody clones: CD45 (BV786 BD HI30), CD3 (APC BD Biosciences 5H9), CD4 (V450 BD Bioscience SK3), CD8 (V500c BD Bioscience SK1), CD14 (FITC BD Bioscience MφP9), sCD19-PE (BioLegend custom conjugation). Cells were then washed twice and resuspended in FACs buffer followed by addition of 7AAD (BioLegend). Live CAR-T cells and non-CAR-T cells were acquired separately using a BD FACSAria utilizing the gating strategy in **Supplementary Fig. 1** with gating based on healthy non-CAR transduced donor PBMC prepared separately for each batch. Samples were resuspended in 0.04% ultrapure bovine serum albumin in PBS at 1000 cells/uL density prior to loading on a 10x Genomics Chromium platform. Infusion product and baseline samples were thawed similarly and combined into batches of up to four samples per 10x channel loaded at a density of 3000 cells/uL. For some infusion product samples with a poor-post thaw viability (<80%), the Miltenyi Dead Cell Removal kit (130-090-101) was used to deplete dead cells prior to scRNAseq.

### Single-cell transcriptome sequencing data generation

Viable cells were resuspended in PBS with 0.04% BSA at a cell concentration of 1000 cells/μL. 40,000 cells were loaded onto a 10x Genomics Chromium instrument (10x Genomics) according to the manufacturer’s instructions. The scRNA-seq libraries were processed using Chromium single cell 5’ library & gel bead kit v2 and coupled scTCR-seq libraries were obtained using Chromium single cell V(D)J enrichment kit (human T cell) (10x Genomics). Quality control for amplified cDNA libraries and final sequencing libraries were performed using Bioanalyzer High Sensitivity DNA Kit (Agilent). Both scRNA-seq and scTCR-seq libraries were normalized to 4nM concentration and pooled in a volume ratio of 4:1. The pooled libraries were sequenced on Illumina NovaSeq S4 platform. The sequencing parameters were: Read 1 of 26bp, Read 2 of 90bp, index 1 of 10bp and index 2 of 10bp.

### Generation of CAR reference

A custom reference was built using GRCh38 and the Ensembl 100 annotation, supplemented with sequences for both the axi-cel and tisa-cel constructs. To determine the sequence of these constructs, we created an initial reference including the sequences of pELPs 19-BB-z^43^ for tisa-cel, and msgv-fmc63-28z^44^ for axi-cel. We then aligned read 2 of four of our infusion product samples (Tisa-R-21, Tisa-N-20, Axi-R-1, Axi-N-14) to this supplemented genome using STAR v2.5.1b. We used the bcftools call tool (v1.11) to identify variants of the true CAR sequence from our initial reference, and the bcftools consensus tool to create an updated fasta file with refined CAR sequences. This was then added to our GRCh38-based reference and prepared with Cellranger mkref (v6.0.1) giving the reference genome used for preprocessing of the full dataset.

### Preprocessing of scRNA-seq GEX data

Raw fastq files for all 10x runs were aligned and quantified using Cellranger v6.0.1 with the expect cells parameter set to 5k per pooled sample (e.g. a run with 5 multiplexed samples was set to 25k) and using the custom reference described above. Gene expression matrices were further filtered for ambient contamination using cellbender v0.2.0.

For all cell multiplexed samples, we ran souporcell v2020.7 to identify genotype clusters using the 1000 genomes common SNP sites. The genotype of every patient at these sites was identified by running bcftools call on the CAR-sorted day 7 samples (since these were not multiplexed). Then the identity of genotype clusters was re-identified by finding the patient with the least discordant genotype calls expected to be in the pool. TCR sequencing data for all runs was processed using cellranger vdj (v6.0.1) and the GRCh38_vdj_v5.0.0 reference package.

### Classification of cell types

An initial clustering of the cells was performed using scanpy^45^ v1.8.1 and a standard single cell workflow. Doublets were filtered by running scrublet^46^ v0.2.1 on each 10x channel individually for PBMC samples. Cells were filtered for a minimum of 200 genes and <15% mitochondrial RNA transcripts. Data was log-normalized as ln(1+10^4^ n_g_/N). Variable genes were selected using scanpy’s highly_variable_genes process with a minimum dispersion of .2 and mean expression values between 0.01 and 3. We further blacklisted from clustering consideration the axi-cel and tisa-cel genes, any TCR or BCR variable genes, as well as sex-biased genes *XIST, RPS4Y1* and *RPS4Y2*. We regressed out the effects of total UMI counts and mitochondrial RNA percentages, and then standardized the expression values with a maximum of 10. We performed PCA, then used harmony to integrate our data batches. Nearest-neighbors were computed using the top 18 harmony-adjusted PCs, and UMAP was run to produce the global visualizations shown.

### Classification of CD4 and CD8 populations

Due to scRNA sparsity CD4 and CD8 T cells cannot be directly classified by gene expression, and clustering at reasonable resolutions did not always cleanly separate these populations. To overcome this, we computed a nearest-neighbor graph using k=100. We then calculated smoothed expression of *CD4* and *CD8A* by taking the mean across the 100 nearest neighbors of every cell. We then classified CD4 and CD8 T cells by gating on these expression values (**Supplementary Fig. 3**).

### Quantification of CD45 isoform expression

Because the important splicing events to distinguish *CD45RA* and *CD45RO* isoforms occur at the 5’ end of the gene (exons 4-7), we were able to develop an isoform quantification model similar to those commonly employed in bulk RNA sequencing^47^. First for every read, we calculated a binary matrix of values *C*_*r,i*_ which are 1 if splicing of read *r* was compatible with each of the six human isoforms *i* of CD45 (RO, RA, RAB, RB, RBC, RABC). We then calculated what the implied fragment length of the sequencing molecule (L_r_^*i*^) would be for each isoform *i* as the distance from the start of the gene to the 3’ end of the read (counting only exons included in isoform *i*). The likelihood of a given read coming from a molecule with a particular isoform *i* could then be modeled as

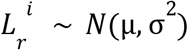

The likelihood of a particular transcript *t* being a certain isoform can be calculated by the product rule over all the reads with that transcript’s UMI

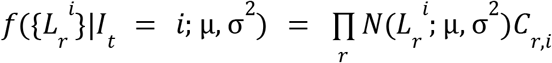

The prior probability of a transcript being of a given class was then modeled with a categorical distribution

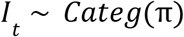

Which allows computation of the posterior probability of a transcript being a particular isoform

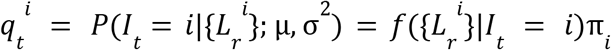

We then applied Expectation-Maximization by iteratively computing the above quantity (E-step), and applying the following update rules

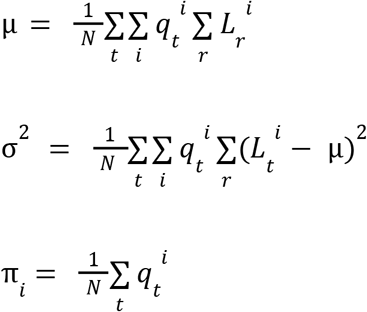

Where *N* is the total number of reads in the sample. In our downstream analyses, we then estimated expression of *CD45RO* and *CD45RA* (the classification antibody which detects isoforms RA, RAB, and RABC) for every cell *c* on the same log-transformed scale as our gene expression data by computing:

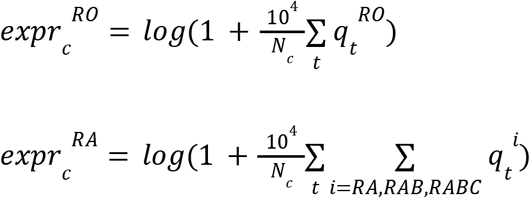

### Supervised classification of T-cell subsets

To label cells, in a fashion consistent with FACS-based literature, we annotated cells based on the expression of commonly used marker genes by thresholding knn-smoothed (k=20) log-normalized expression values as designated in **Supplementary Table 6**. Conventional T-cells were classified by differentiation state as Naive (*CCR7*+*CD45RA*+), CM (*CCR7*+*CD45RA*-), EM (*CCR7*-*CD45RA*-), and TEMRA (*CCR7*-*CD45RA*+). We further distinguished two additional T-cell subsets: T-regs (*FOXP3*+) and CD4+ CTLs (*CD4+NKG7+*).

### Sub-clustering of CAR-T populations

We performed sub-clusterings of CAR-T cells for all eight combinations of CD4/8 subtype, IP/D7 timepoint, and tisa/axi-cel product. Harmony batch correction was used to adjust four samples that had been run without the FACS sorting procedure, using all other samples as a reference. Cells were considered CAR-T if at least a single CAR transcript was observed, or if they shared a TCR sequence with another CAR+ cell. Cells were then processed using the same pipeline outlined above, with the exception that we used fewer (15) PCs and regressed out S and G2M scores. For each of the eight sets, we performed Leiden clustering^48^ to define sub-populations and identified top marker genes with a t-test.

### Testing for compositional changes

We tested for changes in PBMC population frequencies between responders and non-responders by running scCODA^20^ using T-cells as a reference. We fit a model using response and product as predictors, and reported cases found to be credible with >90% inclusion probability (corresponding to a 10% false discovery rate).

When testing overall changes in CAR-T cell populations (**Fig. 4b)**, as a suitable and consistent population to use as a reference is not possible, we used a modified approach. We fit a dirichlet-multinomial model on the cell counts of each patient with parameters ***α***_R_ and ***α***_NR_ for responders and non-responders. We then calculated a log likelihood ratio statistic between the maximum likelihood of this model (L_1_) and that of a null model fixing ***α***_R_=***α***_NR_ (L_0_).

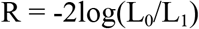

Finally we permuted our responder/non-responder labels 100,000 times and computed a null distribution for R, then used this to calculate an empirical p-value. We reported a Benjamini-Hochberg FDR-corrected q-value, correcting across the 8 categories of CAR-T cells analyzed.

### Pseudobulk differential expression analysis

To perform differential expression while controlling for the dependence structure on the cell level introduced by inter-individual differences, we applied a pseudobulk method. For each gene, we calculated the total number of UMIs in a given cell population of interest by summing across all cells. Cells from different patients, T cell type (Baseline, Infusion-nonCAR, Infusion-CAR, D7-nonCAR, D7-CAR), and CD4/8 subtype were given separate pseudobulk samples, yielding an X by Y matrix of UMI counts. We then analyzed this using a limma ^49^ workflow using multi-level modeling to account for multiple pseudobulk samples coming from the same patient. Counts data was voom-transformed, and sample dependency correlations were computed using the patient ID as a blocking variable. The voom transform was then recomputed using a model with the blocking structure. The limma model was then fitted, and contrasts of interest were compared between populations of interest.

### IP cell type identification

In order to identify common cell populations across different types of CAR-T infusion products (IPs), we re-analyzed just the cells from the IP samples. We used the same workflow as above, however with the difference that we performed Harmony batch correction using the patient ID as a batch variable. This removes the (likely biological) differences between the products, but enabled us to identify common populations between the two, more importantly isolating regulatory T cells in tisa-cel products.

### Independent T-reg validation

We downloaded scRNA-seq gene expression matrices of previously published axi-cel infusion products^9^ from the Gene Expression Omnibus (GEO accession GSE151511). We filtered cells with ≤200 genes and ≥10% mitochondrial RNA. Data was processed using the pipeline described above, using 25 PCs and harmony batch correction on the sample level to allow identification of common populations.

### CAR-Treg validation

#### Lentiviral production

Human embryonic kidney (HEK293T) were purchased from ATCC and expanded in RPMI 1640 media with GlutaMAX and HEPES (ThermoFisher Scientific 72400-120) with 10% FBS (R10). Constructs were synthesized and then cloned into a third-generation lentiviral backbone (GenScript). Replication deficient lentivirus was produced by transfecting plasmids into HEK293T cells. Supernatant was collected at 24 and 48 hours after transfection. Virus was filtered and then concentrated by ultracentrifugation followed by storage at −80C.

#### Bulk T-cell isolation and CAR production

Anonymized human healthy donor leukapheresis products were purchased from the Massachusetts General Hospital blood bank using an IRB-approved protocol. CD3+ T-cells were isolated using a T-cell rosette sep isolation kit (STEM CELL Technologies 15061) and then activated using CD3/CD28 Dynabeads at a 3:1 bead:T-cell ratio (Life Technologies 40203D) in R10 with penicillin/streptomycin (Thermo Fisher Scientific 15140122), and 20IU/mL recombinant human IL-2 (Peprotech 200-02). CARs were transduced with lentivirus at a multiplicity of infection (MOI) of 5 on day 1 and expanded with media doubling and IL-2 replacement every 2-3 days. After one week the CARs underwent magnetic debeading, followed by another week of expansion.

#### Tregulatory cell (Treg) isolation and CAR production

CD4+ T cells were isolated from the autologous donor leukopak (relative to CD3+ cells) using the Rosette-sep Human CD4+ T cell negative selection isolation kit with a Ficoll gradient (STEMCELL Technologies 15062). Isolated CD4+ cells were then enriched for CD25 using CD25-PE antibody staining (BD Bioscience 2A3) and subsequent anti-PE bead selection (Miltenyi Biotec #130-048-801). The resultant two populations (CD4+CD25^high^, and CD4+CD25low) were then stained with CD4 (BV510 BD Biosciences SK3), CD8 (APC-H7 BD Biosciences SK1), CD127 (BV 711 BD Biosciences HIL-7R-M21), and DAPI and sorted on a BD FACS Aria for the following populations DAPI-CD4+CD8-CD25high127- (Tregulatory cells) and DAPI-CD4+,CD8-,CD25- (Control CD4s).

Tregs, CD4-control, and bulk-CD3 cells were activated with anti-CD3/anti-CD28 human Treg expander beads (Thermo Fisher Scientific 1129D) in CTS OpTmizer T cell expansion Serum Free Media (Thermo Fisher Scientific #A1048501) with 2% human serum (Access Cell Culture LLC), 1x GlutaMAX (Thermo Fisher Scientific), 100 U/mL penicillin-streptomycin, and recombinant human IL-2 (300 IU/mL for Tregs and 20 IU/mL for CD4-control). One day after sorting T-cells were transduced at an MOI of 5 with lentivirus. After one week the CARs underwent magnetic debeading, followed by another week of expansion. Transduction efficiency was assessed on day 12-14 after transduction, and non-transduced control cells (Tregs, CD4-control, or bulk-CD3) were added to normalize transduction efficiency across the conditions.

#### FOXP3 staining

Prior to activation, samples were drawn from the CD4+CD25^high^127- (Tregulatory cells), CD4+CD25- (Control CD4s), and CD3+ (Bulk-CD3) groups and stained for FOXP3 (APC Invitrogen PCH101) using the intracellular staining kit (eBioscience 00-5523-00).

#### JeKo-1 lymphoma model

NSG (NOD.Cg-PrkdcscidIl2rgtm1Wjl/SzJ) mice were purchased from Jackson Laboratory and bred in pathogen-free conditions at the MGH Center for Cancer Research. Animals were housed at temperatures of 21.1-24.5C (70-76F), 30-70% humidity, and 12:12 light dark cycles. 6 week old female NSG mice were engrafted intravenously with 10^6^ JeKo-1-click beetle green luciferase (CBG) lymphoma cells (ATCC) on day −7. On day 0, a total of 1×10^6^ fresh CAR-T cells were injected intravenously at the ratios of bulk-CD3, CD4-control, and Tregs as indicated. Mice were imaged biweekly using D-Luciferin (Fisher Scientific) in an Ami HT (Spectral Instruments Imaging). Images were analyzed using Aura version 4.0.0 (Spectral Instruments Imaging) Software.

At the time of euthenasia spleens were collected and sectioned. One section was fixed in 10% neutral buffered formalin, while the remainder was mechanically pulverized and filtered. Cells were stained for CD3 and analyzed via flow cytometry.

Formalin fixed tissue was transferred to 70% ethanol and underwent human CD3 (BD Bioscience UCHT1) staining at the MGH histopathology core. Scale bars were added utilizing Fiji ImageJ (continuously updated, open source, downloaded 2020).

All flow cytometry was analyzed utilizing Flowjo.

Swimmer plot was generated in RStudio (version 2021.09.1) using the package swimplot (version 1.2.0).

### Data visualization

All boxplots presented here show the median, inter-quartile range, and minima/maxima of underlying samples. Overlaid dotplots show individual samples.

### Data availability

Gene expression matrices and raw sequencing data from the scRNA data will be made available upon publication.

## Data Availability

Data produced in the present work will be made publicly available upon publication in a peer-reviewed journal.

## Acknowledgements

We thank Nena Berg, Mary O’Reilly, and the rest of the Broad Institute Patterns team for their contributions to the figure design. This work was partially supported by the Broad/IBM Cancer Resistance Research Project (PIs: G.G., L.P.) and by NIH R01CA252940 (PIs: M.V.M. and G.G.). A special thank you to Dr. Silvia Arrom and Mr. David Oran for their thoughtful donation which helped to support a portion of this work. N.J.H. was partially funded by the Landry Cancer Biology Consortium fellowship. S.L. is supported by the NCI Research Specialist Award (R50CA251956). R.C.L. is supported by NIH T32 AI007529. Biorender.com was utilized for a portion of some figures.

## Figures

**Supplementary Figure 1.**
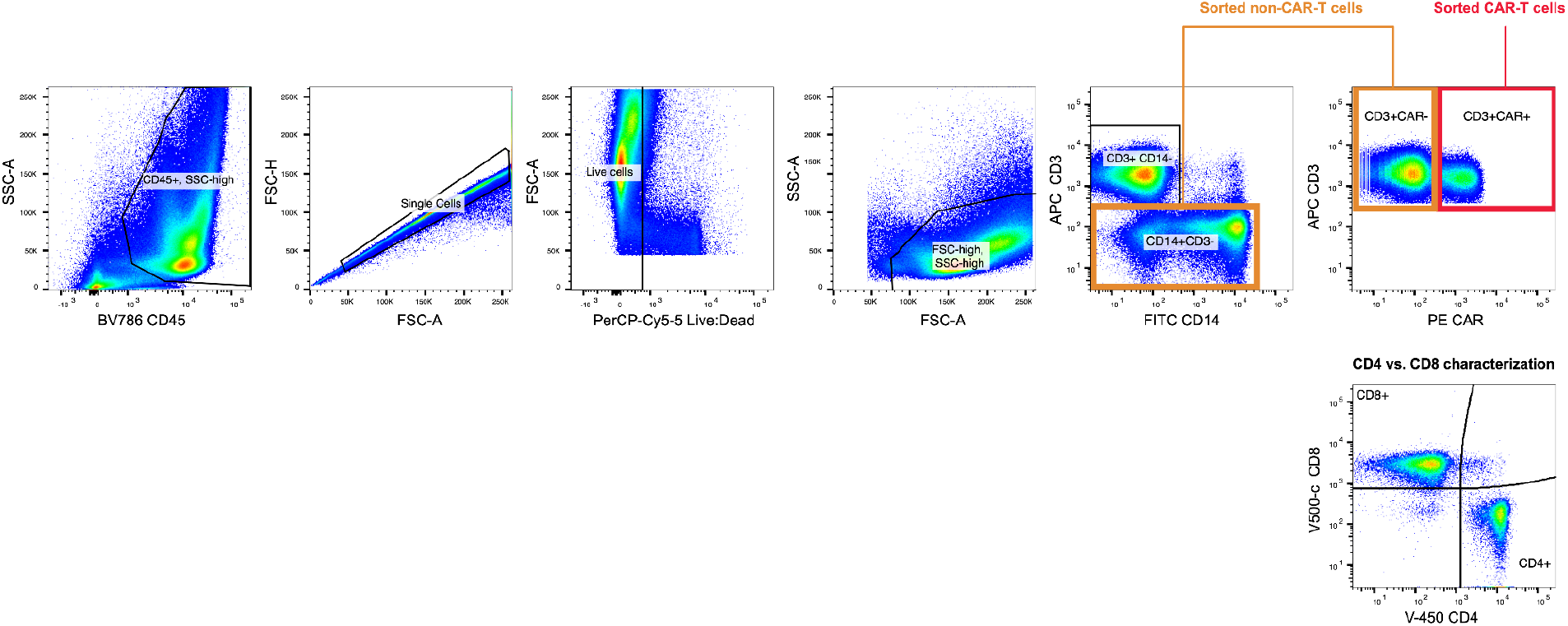
Flow cytometric gating strategy for downstream scRNAseq analysis. Live CD14-CD3+CAR+ cells were isolated as CAR-T cells while CD3+CAR-cells were combined with CD14+CD3-cells as non-CAR-T cells for subsequent scRNAseq workflows.

**Supplementary Figure 2.**
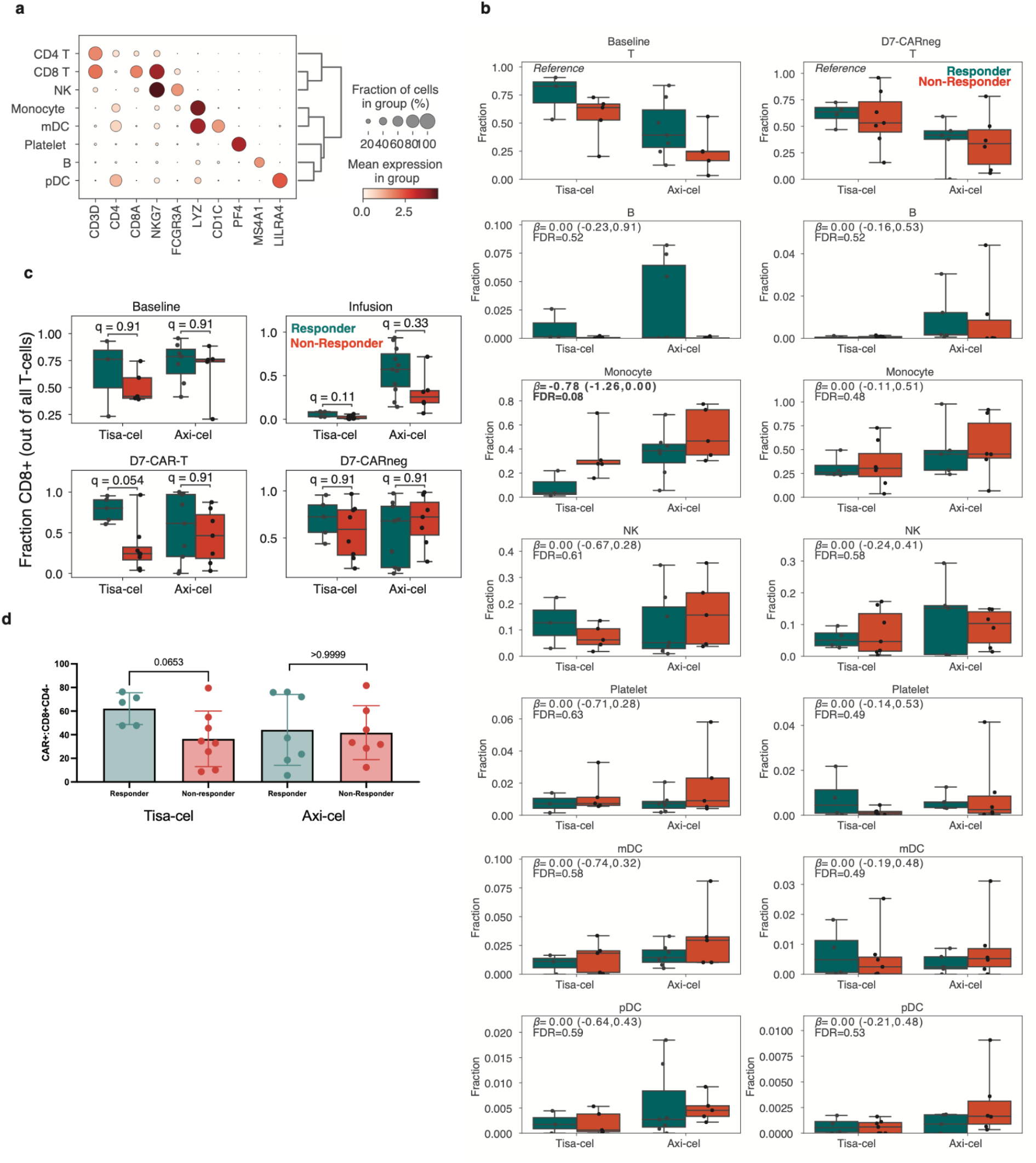
PBMC cell composition differences between responders and non-responders pre- and post-infusion. **a**, Selected marker genes for PBMC cell population identification. **b**, Boxplots of the cell type frequencies for each cell type stratified by product and response. The final cell type coefficient (with its posterior 95% high density interval) and FDR value (one minus the inclusion probability) estimated by scCODA are shown. **c**, Fraction of cells which were CD8+CD4-at baseline, in IPs, and at day 7 in CAR+ and CAR-populations. **d**, Flow-cytometric measurements for fraction of CAR positive cells which were CD8+CD4-at day 7. Error bars represent standard deviation. P-values represent two-tailed Mann -Whitney U tests.

**Supplementary Figure 3.**
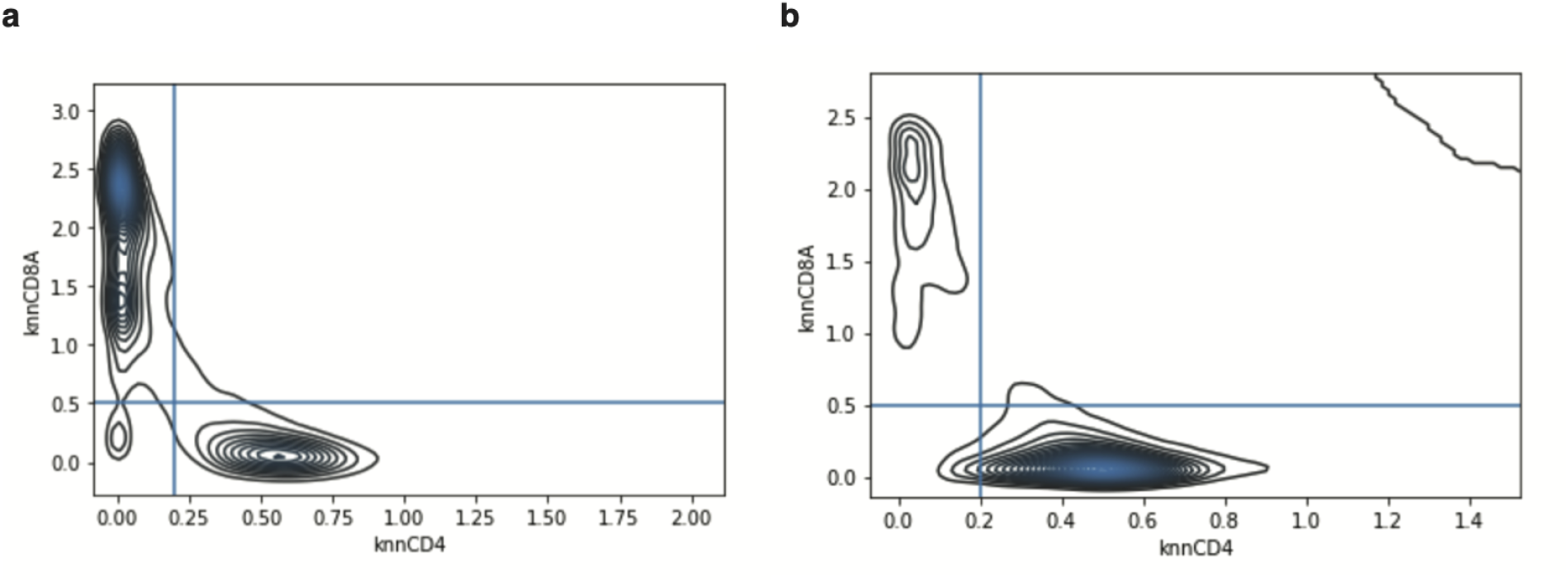
Classification of CD4 and CD8 T-cells. Kernel density estimate plots of knn-smoothed (k=100) CD4 and CD8A expression across cells at **a**, day 7 and **b**, in the IP. Lines are drawn for thresholds used for classification.

**Supplementary Figure 4.**
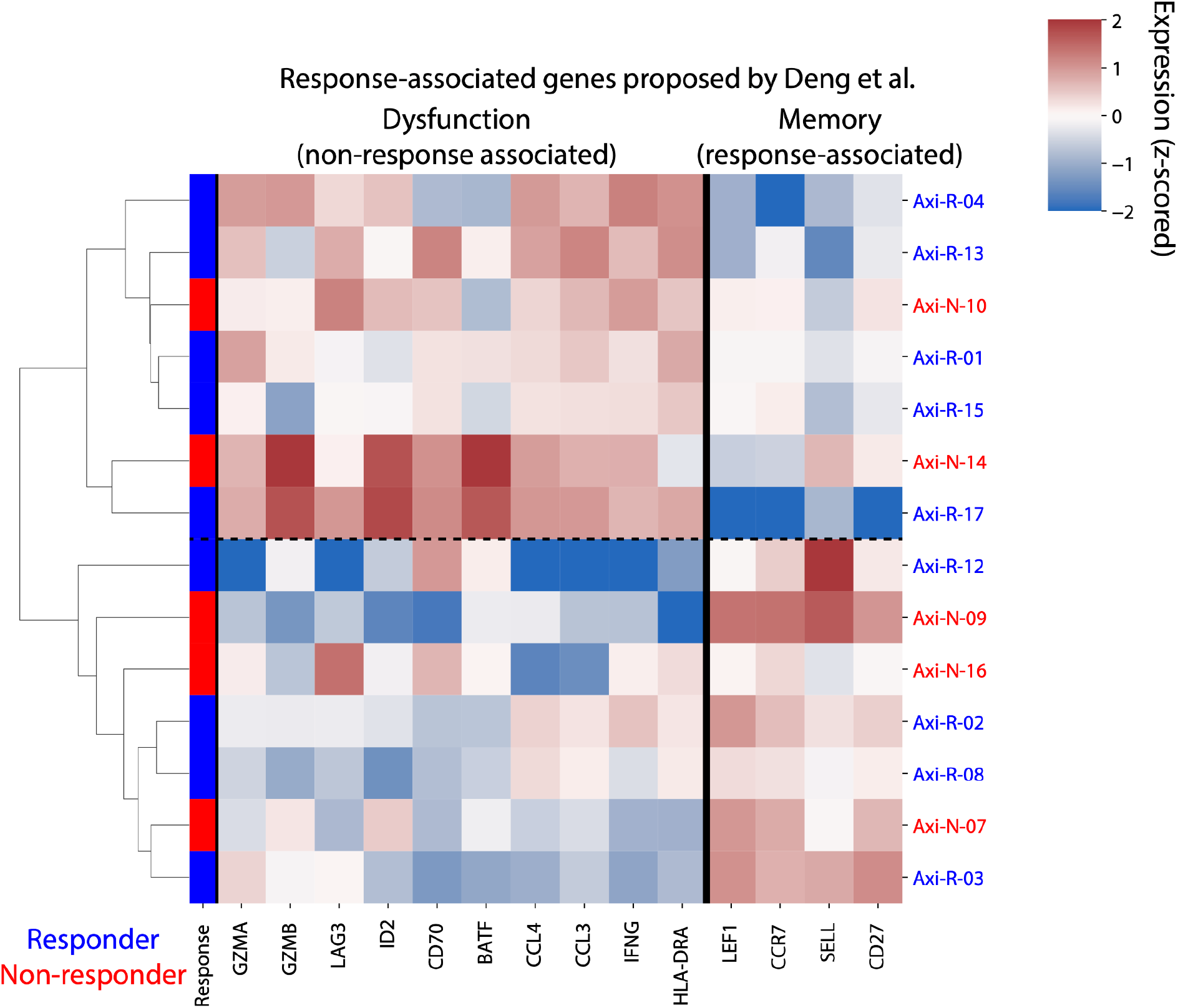
Expression of previously proposed axi-cel response genes. Pseudobulk expression (z-scored log transcripts per million) of CD8+ CAR-T axi-cel IP cells for genes proposed to be response-associated by Deng et al. Fisher Exact test for association between response and denoted two clusters driven by putative memory- and dysfunction-associated genes is p=1.

**Supplementary Figure 5.**
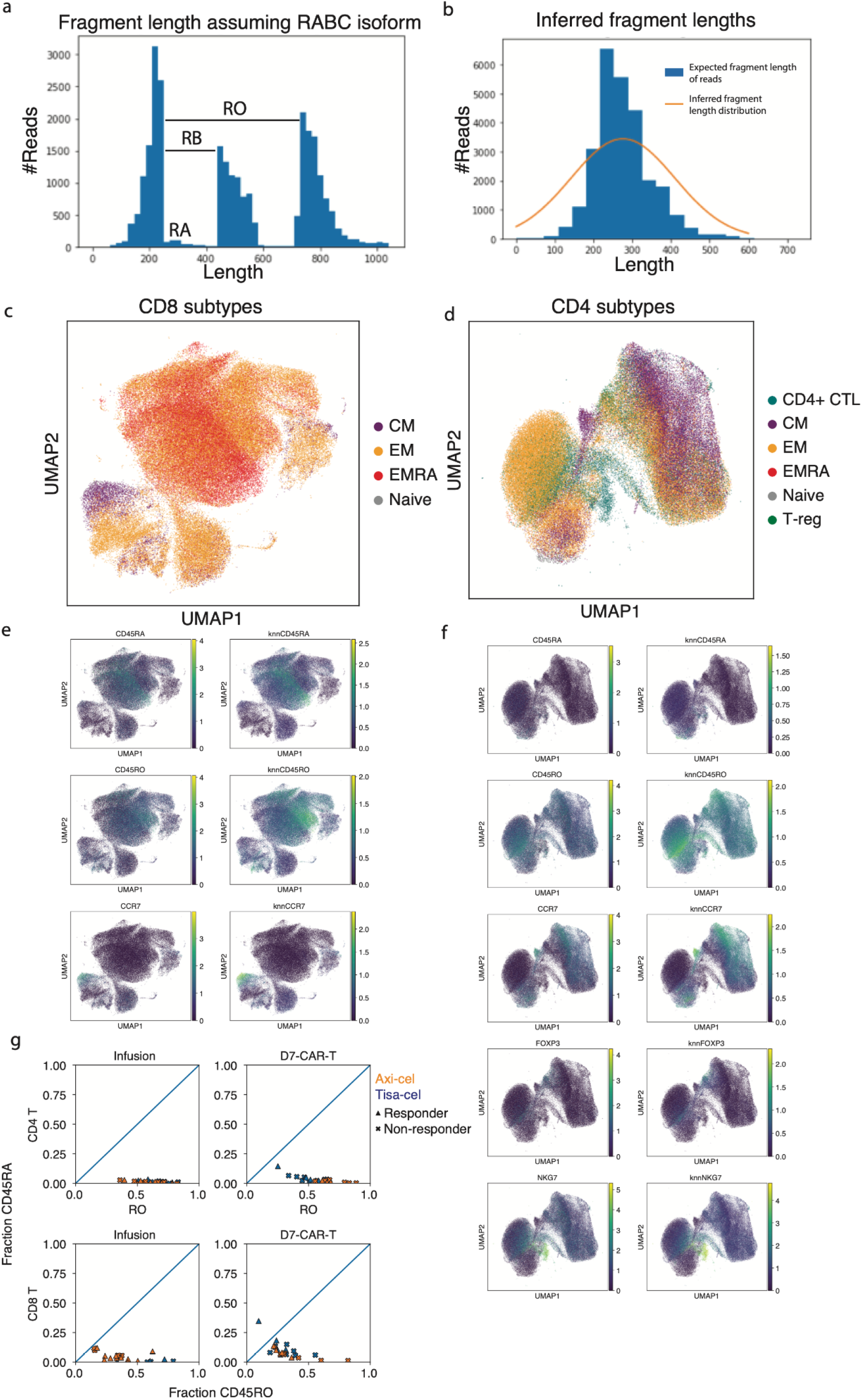
Classification of T cell subtypes. **a**, Illustration of signal used by CD45 isoform detection model. For each read in an illustrative sample, a histogram of the fragment length assuming no splicing (RABC isoform) is shown. The distribution of reads beyond exon 3 become shifted if they come from an isoform lacking an upstream exon. **b**, A histogram of the expected fragment length after inference of each read is shown for the same sample in blue. The gaussian distribution modeling fragment lengths inferred by EM is plotted in orange. **c,d**, UMAP representations of all CD8 and CD4 T cells colored by their subtype assignment. **e,f**, Expression of marker genes used for subtype assignment. For each a knn-smoothed (k=25) estimate is shown (right) alongside the raw measurements (left). **g**, Estimated fractions of CD45 isoforms that are RO (x-axis) or RA (y-axis) in CAR-T cells from infusion products and at day 7.

**Supplementary Figure 6.**
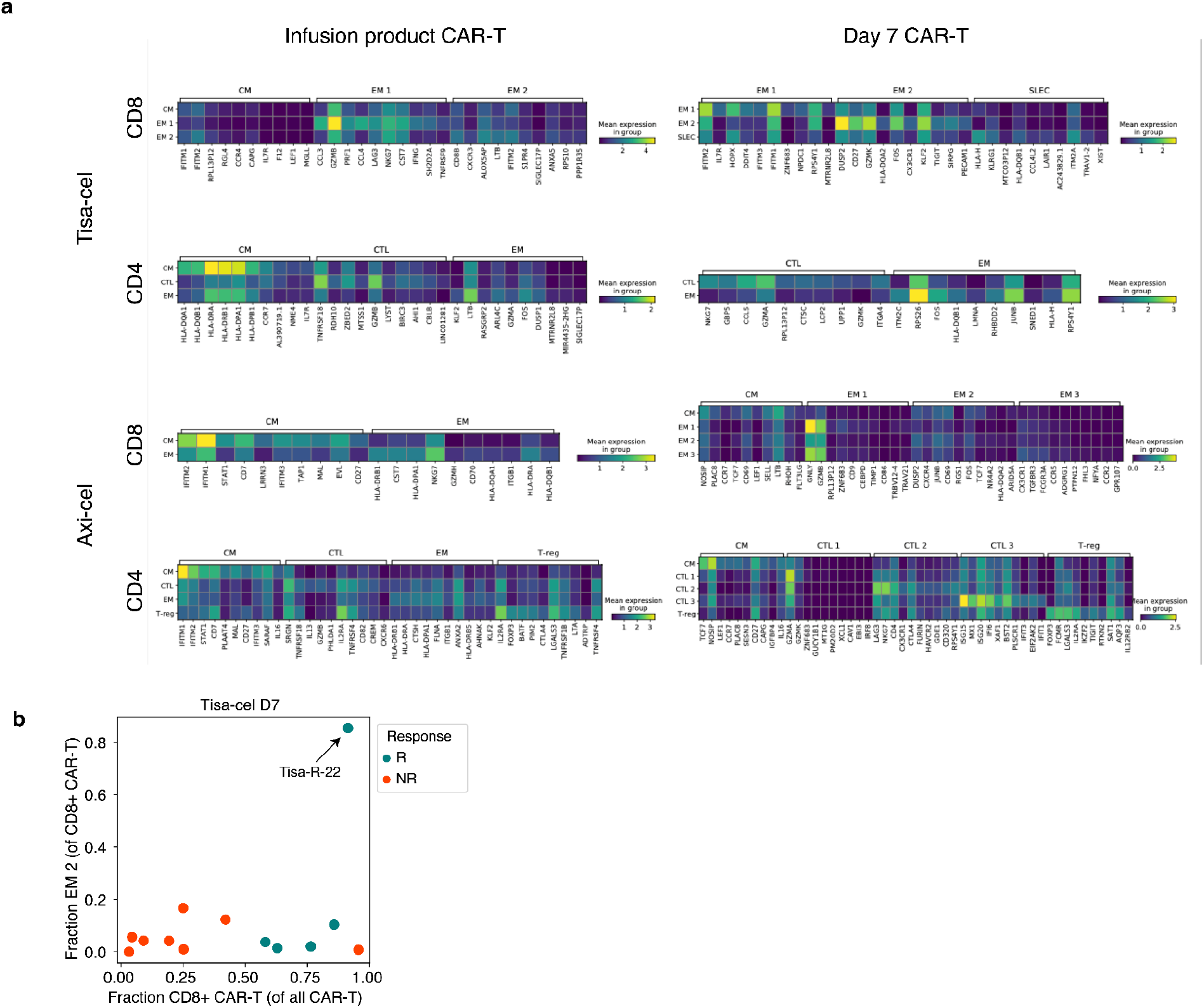
CAR-T subcluster markers and associations. **a**, Top differentially expressed genes in each CAR-T subcluster, as determined by a t-test. The expression is shown for the top 10 marker genes of each cluster displayed in Figure 4a. **b**, Demonstration of unique tisa-cel responder with CD8+ cells in cluster EM 2. Shown is a scatter plot of the fraction of CD8+ cells in cluster EM 2 (y-axis) vs the fraction of all CAR-T cells that are CD8+ (x-axis).

**Supplementary Figure 7.**
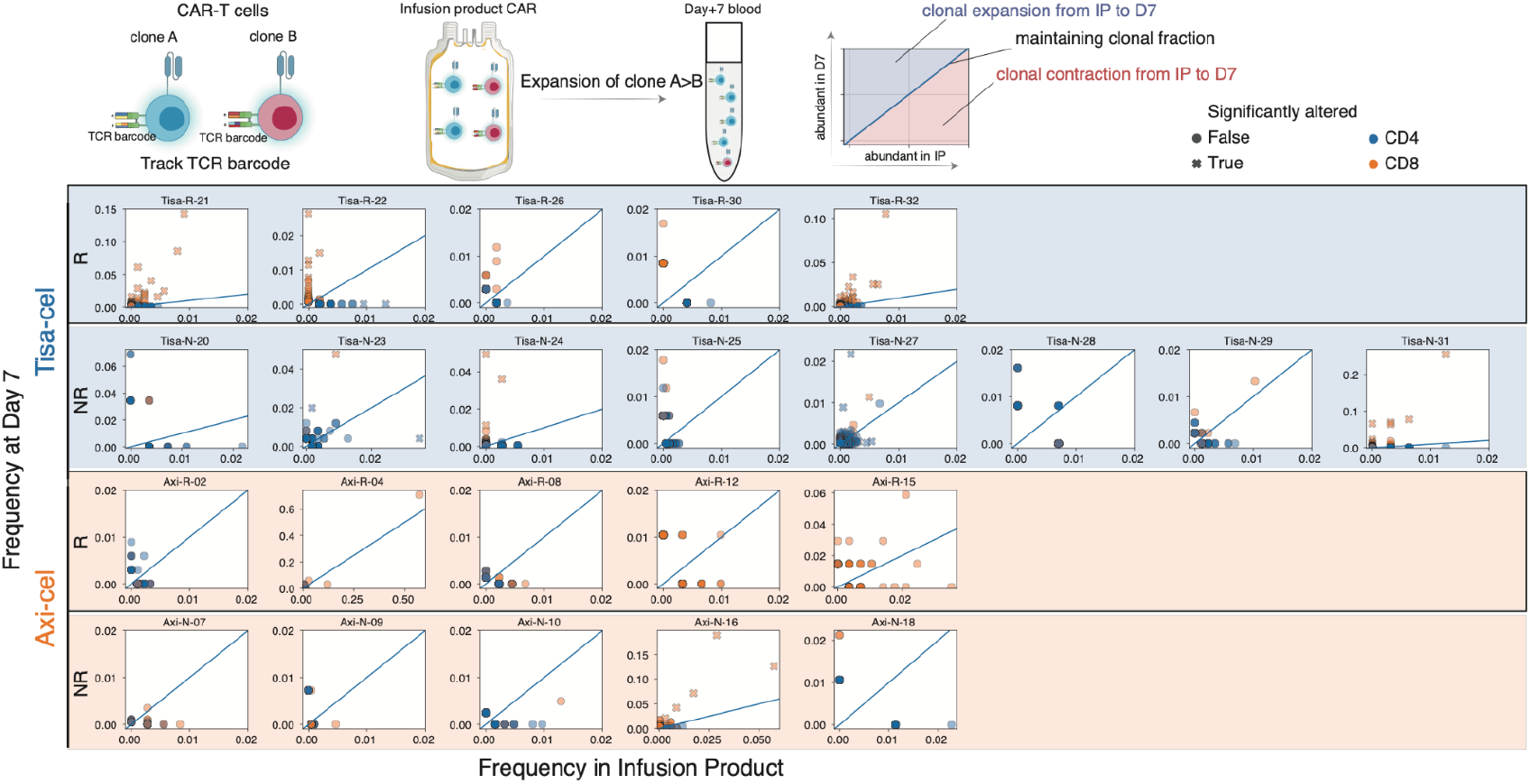
Changes in individual TCR clone frequencies per patient. For each patient with at least 25 T cells in both the IP and at day 7, individual TCR clones are plotted by their frequency in the IP (x-axis) and at day 7 (y-axis). Clones are colored by whether they are CD8+ or CD4+, and denoted with an “x” if the two timepoint frequencies are significantly different by a fisher exact test p<0.05.

**Supplementary Figure 8.**
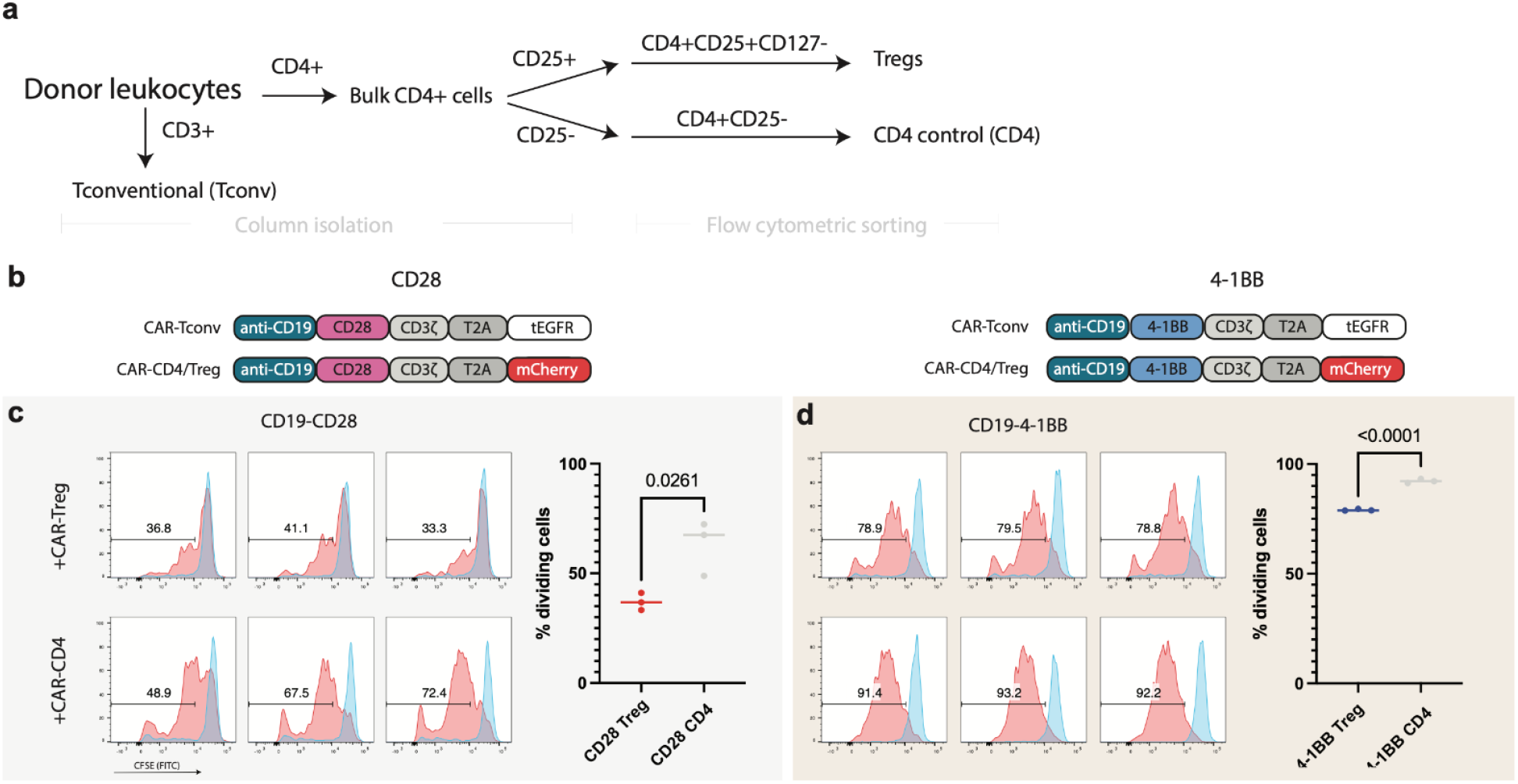
Treg isolation, CAR-Treg generation, and in vitro suppression. **a**. Schematic for T-reg and CD4 control population isolation from healthy donor PBMC. **b**. CAR constructs used to identify CAR-Treg/CD4-CAR cells from CAR-Tconv. **c**. CFSE staining of CAR-Tconv cells co-cultured with either 25% CAR-Tregs or CD4-CAR control cells and stimulated at a 1:1 ratio with Jeko tumor targets at 72 hours. Dividing cells (red) are identified relative to unstimulated condition (blue). Each histogram represents an individual replicate, summarized in the plot on the right. P-value represents two-tailed unpaired t-test.

**Supplementary Figure 9.**
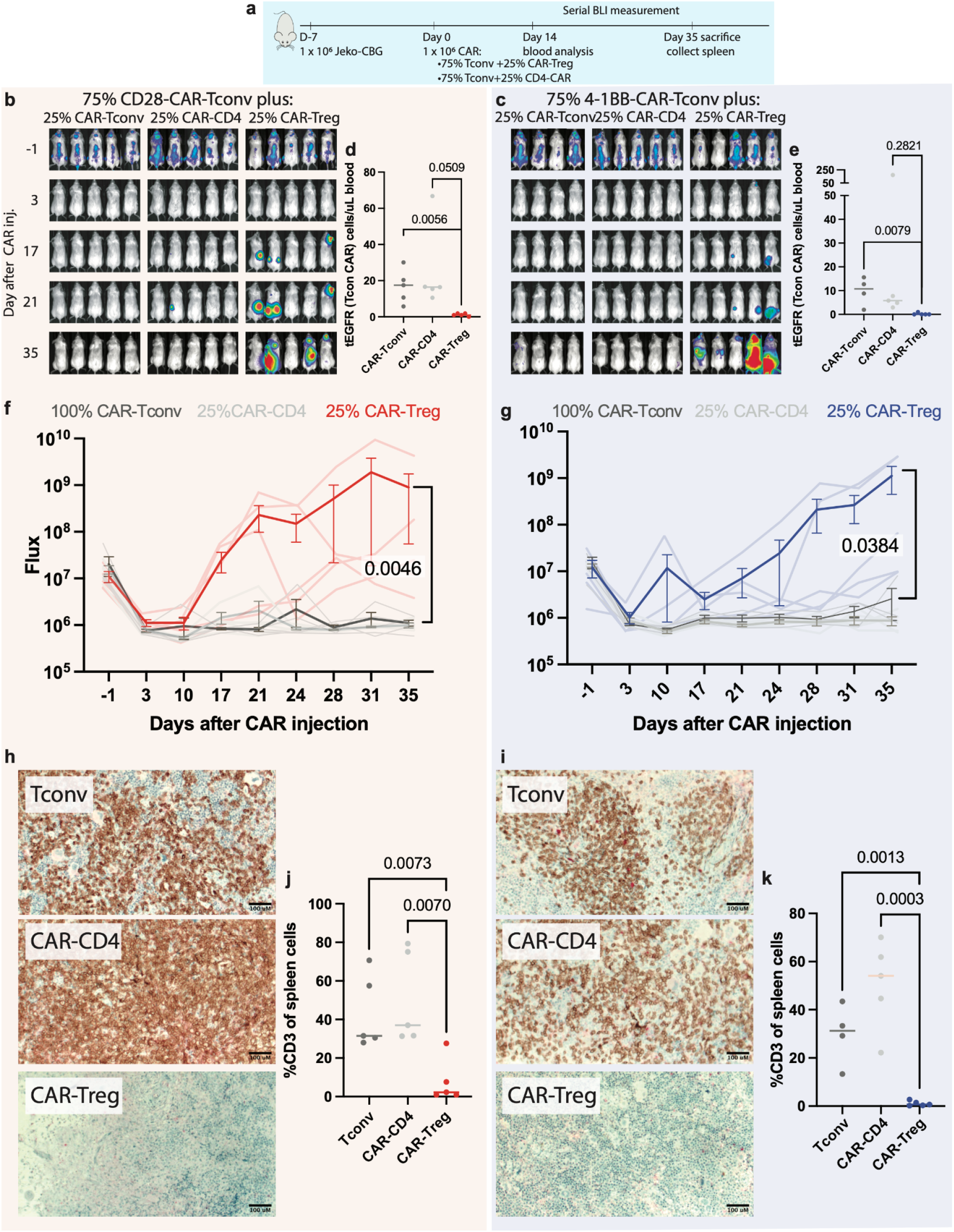
Mediation of relapse by CAR-Tregs at 25%via *in vivo* validation experiments. **a**, NSG mice were injected with 1 × 10^6^ Jeko-CBG lymphoma cells on day −7. On day 0 mice were injected with 1 × 10^6^ CAR-T cells representing 100% CAR-T convs or 75% CAR-Tconvs with either 25% CAR-Tregs or 25% CD4-CAR-T control cells. Experiment performed with CD19-CD28 (left) or CD19-4-1BB (right) constructs. **b,c** Time course tumor radiance (photons/sec/cm2/sr). **d,e** Flow cytometric quantification of CAR-Tconv day 14 after CAR injection. P-value represents two-tailed unpaired t-test. **f,g** Time course flux (photons/s). Mean ± SEM overlayed on individual subject curves. P-value represents the result of two-way ANOVA.. **h,i** Representative immunohistochemical staining for human CD3 in the spleen. **j,k** Flow cytometric quantification of CD3 cells from the spleens of the indicated conditions. P-value represents two-tailed unpaired t-test.

**Supplementary Figure 10.**
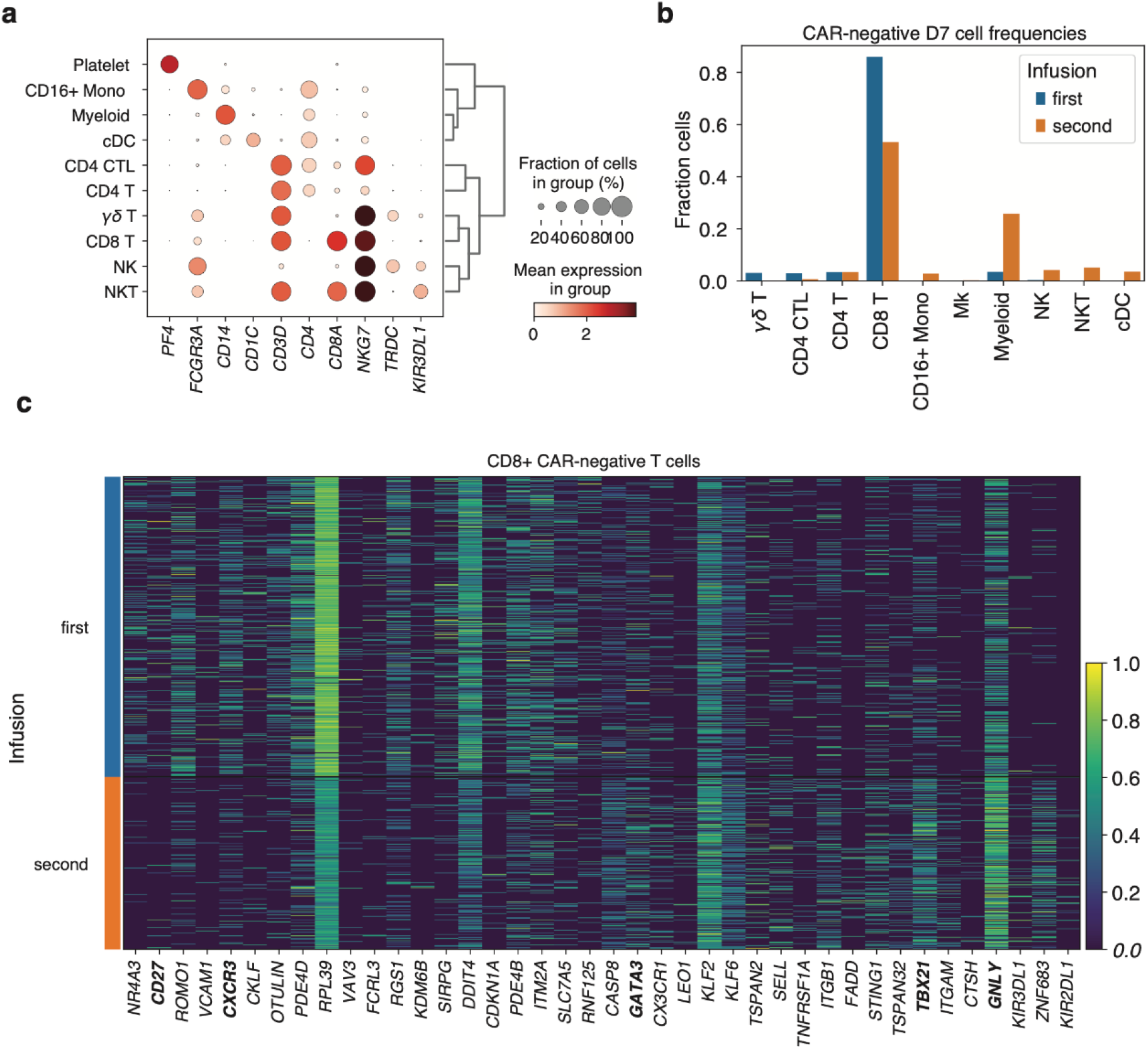
Changes in PBMC populations in patient treated with second infusion of tisa-cel. **a**, Expression of marker genes for PBMC cell type classification. **b**, Fractions of day 7 CAR-negative cells falling into each cell type cluster, stratified by first and second infusion. **c**, Top genes differentially expressed (by Mann-Whitney U test) comparing CD8+ CAR-negative T cells between the first and second treatments at day 7.

**Supplementary Figure 11.**
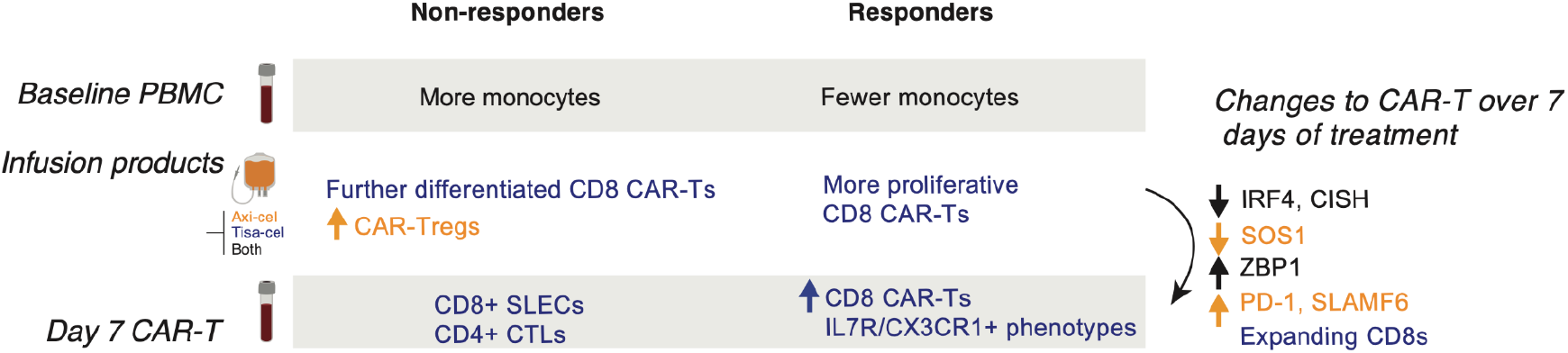
Summary of cellular and transcriptomic changes associated with clinical outcome and timepoint.

## Supplementary tables

**Supplementary Table 1. Characteristics of patients included in this study**

**Supplementary Table 2. List of single-cell RNA samples and quality metrics**

**Supplementary Table 3. Genes differentiation upregulated in CAR+ vs CAR-cells**

**Supplementary Table 4. Genes upregulated at day 7 compared to IP**

**Supplementary Table 5. Genes differentially expressed in cells of responders vs non-responders**

**Supplementary Table 6. Marker genes used for supervised T-cell classification**

